# The National Registry of Rare Kidney Diseases (RaDaR): Description, recruitment, and cross-sectional analyses of 25,880 adults and children with rare kidney diseases in the UK

**DOI:** 10.1101/2023.09.24.23296009

**Authors:** Katie Wong, David Pitcher, Fiona Braddon, Lewis Downward, Retha Steenkamp, Nicholas Annear, Jonathan Barratt, Coralie Bingham, Richard J. Coward, Tina Chrysochou, David Game, Sian Griffin, Matt Hall, Sally Johnson, Durga Kanigicherla, Fiona Karet Frankl, David Kavanagh, Larissa Kerecuk, Eamonn R. Maher, Shabbir Moochhala, Jenny Pinney, John A. Sayer, Roslyn Simms, Smeeta Sinha, Shalabh Srivastava, Frederick W. K. Tam, Kay Thomas, A. Neil Turner, Stephen B. Walsh, Aoife Waters, Patricia Wilson, Edwin Wong, RaDaR consortium, Karla Therese L. Sy, Kui Huang, Jamie Ye, Dorothea Nitsch, Moin Saleem, Detlef Bockenhauer, Kate Bramham, Daniel P. Gale

## Abstract

Rare kidney diseases are not well characterised, despite making a significant contribution to the burden of kidney disease globally. The National Registry of Rare Kidney Diseases (RaDaR) collects longitudinal disease and treatment-related data from people living with rare kidney diseases across the UK, and is the largest rare kidney disease registry in the world. We present the clinical demographics and renal function of 25,880 prevalent patients and evaluate for any potential recruitment bias to RaDaR.

RaDaR has automatic linkage with the UK Renal Registry (UKRR, with which all UK patients receiving Kidney Replacement Therapy (KRT) are registered). To assess for recruitment bias to RaDaR, ethnicity and socioeconomic status of 1) prevalent RaDaR patients receiving KRT were compared with patients with eligible rare disease diagnoses receiving KRT in the UKRR 2) patients recruited to RaDaR and all eligible unrecruited patients at two renal centres were compared 3) the age-stratified ethnicity distribution of RaDaR patients with Autosomal Dominant Polycystic Kidney Disease (ADPKD) was compared to the English Census.

We found evidence of some disparities in ethnicity and social deprivation in recruitment to RaDaR, however these were not consistent across all comparisons. Predominant rare kidney diseases in adults were ADPKD (29.2%), Vasculitis (15.8%) and IgA nephropathy (15.7%), compared to Idiopathic nephrotic syndrome (43.6%), Vasculitis (10.8%) and Alport Syndrome (5.9%) in children. Compared with either adults recruited to RaDaR or the English population, children recruited to RaDaR were more likely to be of Asian ethnicity and live in more socially deprived areas.

**Lay Summary:** Rare kidney diseases make a significant contribution to the number of people living with kidney disease globally: >25% of adults and >50% of children with kidney failure have a rare disease. However, there is a lack of high-quality published data on how these conditions present, and in which patient groups. Patients often face delays in diagnosis and lack of reliable information on their condition once diagnosed. The UK National Registry of Rare Kidney Diseases (RaDaR) was formed in 2010 to address this knowledge gap. It collects long-term data for UK patients with rare kidney conditions, and is the largest rare kidney disease registry in the world. Here, we present information about 25,880 adults and children recruited to RaDaR, including ethnicity, socioeconomic status, and kidney function, and investigate whether there is any bias in recruitment to RaDaR. To our knowledge, this is the largest epidemiological description of rare kidney diseases worldwide.

## Introduction

A rare disease is defined in Europe as a condition affecting less than 1 in 2000 people ^1^, and in the USA as affecting fewer than 200,000 individuals in the country^2^. Rare kidney diseases make a significant contribution to the burden of kidney disease in the UK and globally: at least 25% of adults and over 50% of children receiving kidney replacement therapy (KRT) have a rare disease^3^ with ’glomerulonephritis’ the single commonest category of Primary Renal Disease among UK patients receiving KRT^4^.

Small patient numbers can result in challenges in clinical management and research in rare diseases: lack of clinical experience, even in large academic centres can lead to delays or errors in diagnosis and treatment of rare diseases; and low disease incidence alongside underdiagnosis can make identification of patients eligible for clinical trials and observational studies challenging. Adequate patient numbers for meaningful analysis may only be achieved through collaboration between multiple large renal centres, associated with considerable administrative burden^5^.

Kidney disorders can cause multi-system dysfunction and may require complex multidisciplinary care at different specialist centres. Advances in KRT have led to people with rare kidney disorders surviving for decades with Kidney Failure (KF)^1^ so the requirement for long-term follow-up data is paramount. For children with rare kidney diseases, life-time follow-up across different specialist paediatric and adult healthcare centres across different regions may be needed, leading to fragmentation of records across multiple databases, systems, and health care providers which is challenging to access for research. Rare kidney disorders are therefore frequently poorly characterized, lacking published data on the prevalence rates, determinants, distribution, and long-term outcomes of these diseases.

The National Rare Kidney Disease Registry (RaDaR), set-up in 2010 by the UK Kidney Association (UKKA) with funding from the Medical Research Council, Kidney Care UK and Kidney Research UK, was designed to address these challenges, by collecting longitudinal data for UK adults and children with rare kidney diseases. Uniquely embedded with the publicly funded National Health Service (NHS) to which all UK residents have free access, RaDaR is hosted by the UK Renal Registry (UKRR) and has UK-wide ethical approval as a research registry, enabling automated collection of retrospective and prospective data for patients across multiple healthcare systems and regions. The aims of RaDaR include to i) better understand the natural history of rare kidney diseases; ii) assess long-term effects of therapies; iii) identify cohorts eligible for clinical research; iv) provide infrastructure for individual rare disease studies and sub-registries.

RaDaR is the largest rare kidney disease registry worldwide. Here we describe the set-up and data flow into RaDaR, and present cross-sectional analyses of 25,880 prevalent patients and minimum point prevalence estimates for 21 rare kidney diseases in the UK.

## Materials and methods

### Structure of RaDaR

Recruitment and data transfer are summarised in Figure 1. All participants sign a paper or e-consent form, consenting to access and analysis of retrospective and prospective clinical data, and in most cases to sharing of their data from other databases, studies and registries, and to be contacted for future research studies they may be eligible for. All data are held centrally in a Structured Query Language database at the UKRR. NIHR (National Institute for Health and Care Research) infrastructure and research nurse supports NHS sites to manually enter a minimal set of mandatory fields at time of recruitment to RaDaR; this infrastructure has also supported other national research programmes such as the RECOVERY^6^ trial. Manual data entry is automatically checked using defined ranges to identify implausible data.

**Figure 1:**
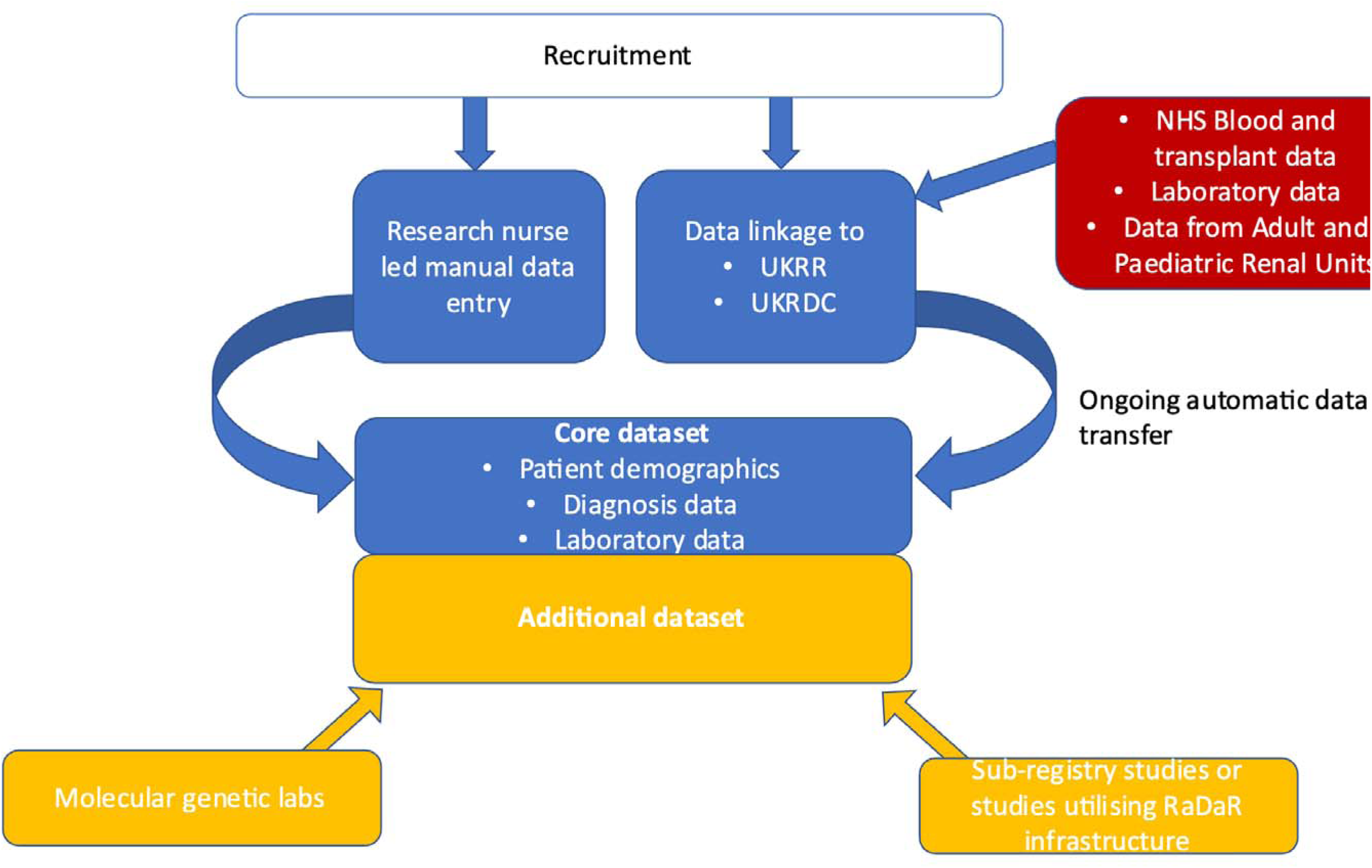
Recruitment and data flow to RaDaR

Data on all patients recruited to RaDaR were extracted on July 25^th^ 2022.

### Rare disease groups (RDGS)

Participants are recruited into 29 Rare Disease Groups (RDGs), which may comprise a single disease entity or groups of renal diagnoses. Full eligibility criteria for each RDG are in Supplementary Appendix 2 and available online at https://ukkidney.org/sites/renal.org/files/radar/Inclusion-Exclusion_april_2021_v22.pdf.

Data for RDGs with > 85 patients recruited are presented: Autosomal Dominant Polycystic Kidney Disease (ADPKD), Autosomal Dominant Tubulointerstitial Kidney Disease (ADTKD), Atypical Haemolytic Uraemic Syndrome (aHUS), Alport Syndrome (AS), Autosomal Recessive Polycystic Kidney Disease and Nephronophthisis (ARPKD/NPHP), Hepatocyte Nuclear Factor-1 Beta (*HNF1B*) mutations, IgA nephropathy, Idiopathic Nephrotic Syndrome (INS), Membranous Nephropathy, Monoclonal Gammopathy of Renal Significance (MGRS), Membranoproliferative Glomerulonephritis and C3 Glomerulopathy (MPGN/C3GN), Pregnancy, Inherited Renal Cancers, Retroperitoneal Fibrosis, Shiga Toxin/verotoxin-producing *Escherichia Coli* associated Haemolytic Uraemic Syndrome (STEC-HUS), Tuberous Sclerosis Complex (TSC), Vasculitis, Cystinosis, Cystinuria, Primary Hyperoxaluria and Tubulopathies.

Data from individuals with AS with a clinical diagnosis of X-linked AS or Thin Basement Membrane Nephropathy (TBMN) are reported with results for males and females with X-Linked AS, and individuals with TBMN presented separately. Patients with two *COL4A3* or *COL4A4* pathogenic variants (autosomal recessive AS) have been excluded due to small sample size.

Results for the Vasculitis RDG are presented stratified into ANCA-associated vasculitis, Anti-GBM disease and Other Vasculitides (including large vessel and IgA vasculitis). The INS cohort is presented stratified into patients with a diagnosis of either Steroid Sensitive Nephrotic Syndrome or Minimal Change Disease (SSNS/MCD); Steroid Resistant Nephrotic Syndrome, Congenital Nephrotic Syndrome or Focal Segmental Glomerulosclerosis (SRNS/FSGS); or INS-unspecified (patients without a confirmed diagnosis of SSNS/MCS or SRNS/FSGS).

### Data linkage

RaDaR is linked with the UKRR for data on KRT initiation and death, which receives data from NHS Blood and Transplant for transplantation events. Rates of KRT data received from the UKRR are correct as of 1st January 2022. Routine laboratory data are extracted via automated feed either directly from renal unit IT systems or via the UK Renal Data Collaboration (UKRDC).

### Demographic data

Self-reported patient ethnicity (Office of National Statistics (ONS) census categories^7^) is entered manually by a research nurse at time of recruitment or populated from existing clinical data provided by the UKRDC. Sex is recorded according to clinical record. Postcodes (zipcodes) were used to derive Index of Multiple Deprivation (IMD) scores as an area level measure of socioeconomic status (SES). IMD is a measure of relative deprivation for small areas within a country from most deprived to least deprived (1 = most deprived). Each country in the UK (England^8^, Wales^9^, Scotland^10^ and Northern Ireland^11^) has an IMD. These can then be categorised into country-specific quintiles within each country (Quintile 1= most deprived, Quintile 5= least deprived).

### Renal function

For patients not receiving KRT, eGFR was calculated using CKD-EPI Cr equation without race adjustment (2009)^12^ or Schwartz equation for patients ≤16 years old.

### Missing data

Available data are presented for each variable and patterns for missing data explored and proportions presented in Supplementary Tables 1-3.

### Small number suppression

Where a risk of re-identification of participants has been identified, groups with small numbers have been aggregated into larger groups and tabulated data has been structured not to report fewer than 6 participants per cell where possible. Where cells contained ≤6 counts, this cell has been suppressed. To avoid possibility of calculation of suppressed counts, corresponding cells are rounded to the nearest 5, in accordance with NHS Digital guidance^13^.

### Minimum point prevalence estimates

UK wide RaDaR point prevalence estimates were calculated using patient numbers for each RDG, and stratified by sex, using ONS UK population data^14^ and are presented per 100,000 population. Prevalence estimates were also calculated for each RDG for each UK Health Board, and maximum estimated rate for each RDG are presented. Due to the nature of recruitment to RaDaR, which requires informed consent from participants, these estimated UK-wide rates will under-estimate the true rate of rare kidney diseases but could be interpreted as minimum possible rates.

### Statistical analyses

Baseline characteristics are presented as frequencies (%) for categorical data and medians (IQR) for continuous data. Chi-square or Fishers exact tests were used to compare categorical variables. Statistical analyses were performed using STATA Release 17 and SAS version 9.4.

### Ethnicity and Social Deprivation comparisons

For each RDG, the proportion of patients in each ethnic group and each IMD Quintile was compared to the overall RaDaR proportion, excluding that RDG. As ethnic and IMD Quintile distributions differed between adult and paediatric populations, these analyses were also performed stratified by age category.

### Recruitment comparisons

Primary renal diagnosis (PRD) is recorded in the UKRR using ERA-EDTA (European Renal Association-European Dialysis and Transplant Association) codes^15^ and in UK renal IT systems either using PRD codes or free-text. PRD codes and a list of search terms specific to each RDG were decided with agreement from RDG leads (clinicians with expertise in that rare kidney disease), to generate an overall list of PRD codes and keywords for RaDaR diagnoses (Supplementary Table 4).

To assess whether some RDGs had recruited a greater proportion of the total eligible patients in the UK than others, living patients in the UKRR receiving KRT who are eligible for RaDaR based on EDTA codes were stratified into their potential RDGs, and the percentage in each RDG compared to the percentage of living patients in RaDaR receiving KRT.

To assess whether there has been ethnic or SES recruitment bias to RaDaR, three methods were used: 1) Ethnicity and SES of all prevalent RaDaR patients who had reached KF were compared with patients with a rare kidney diagnosis in the UKRR 2) Patients recruited to RaDaR from two large UK renal centres were compared with all unrecruited patients with a RaDaR eligible diagnosis at those centres 3) the age-stratified ethnicity distribution of England according to the 2011 UK census was compared with the ethnicity of prevalent English RaDaR patients with ADPKD. Patients from Scotland, Wales and Northern Ireland were excluded from comparisons with the 2011 UK census due to lack of available data regarding age-stratified ethnicity from the ONS for those nations. English nationality was determined by a home address with an English postcode. More detailed information about these comparisons is presented in the Supplementary Methods.

The RaDaR database has approval for research studies from the NHS South-West-Central Bristol Research Ethics Committee (19/SW/0173). The report was written with reference to the Strengthening the Reporting of Observation Studies in Epidemiology (STROBE) statement^16^

## Results

As of July 2022, RaDaR recruits patients from 108 NHS sites (96 adult and 12 paediatric) across England (n=91), Scotland (n=9), Wales (n=3) and Northern Ireland (n=5). Most patients have been recruited from English renal units (n=23,776, 92%). Data from RDGs with ≥85 patients recruited are presented.

### Clinical demographics of the RaDaR Patient Population

Clinical characteristics of 25,880 prevalent patients in RaDaR on 25^th^ July 2022 are displayed in Table 1. 2,957 patients are now deceased. 125 patients (0.5%) have more than one diagnosis recorded; the majority of these are in the pregnancy RDG (115/125, 92%).

**Table 1:**
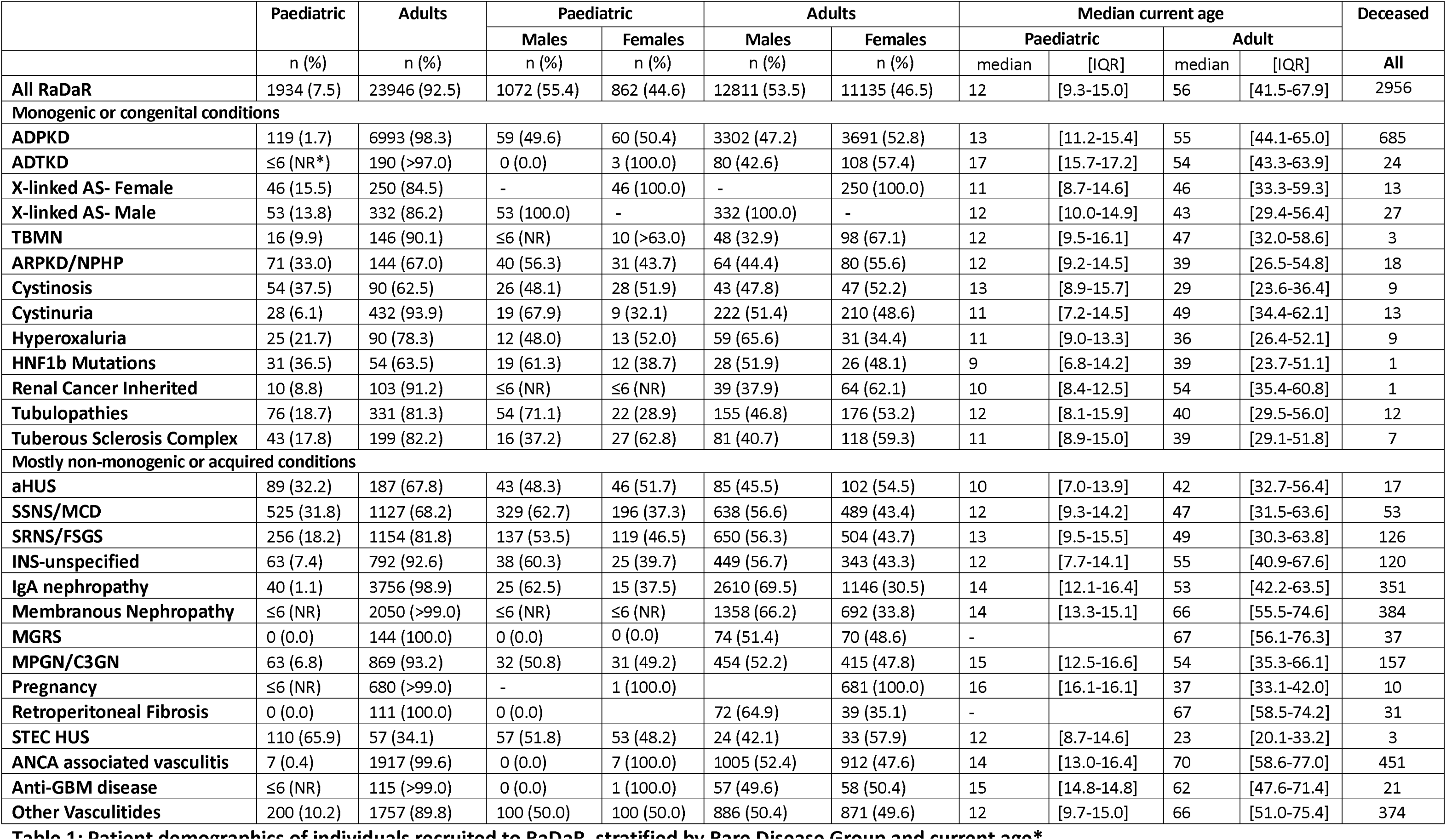
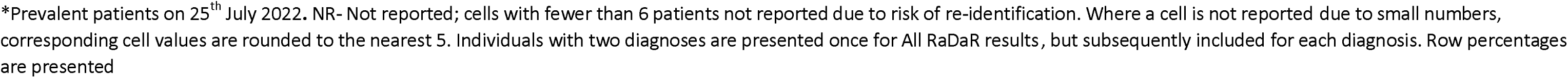
Patient demographics of individuals recruited to RaDaR, stratified by Rare Disease Group and current age*.

|  | Paediatric | Adults | Paediatric |  | Adults |  | Median current age |  |  |  | Deceased |
| --- | --- | --- | --- | --- | --- | --- | --- | --- | --- | --- | --- |
|  |  |  | Males | Females | Males | Females | Paediatric |  | Adult |  |  |
|  | n (%) | n (%) | n (%) | n (%) | n (%) | n (%) | median | [IQR] | median | [IQR] | All |
| All RaDaR | 1934 (7.5) | 23946 (92.5) | 1072 (55.4) | 862 (44.6) | 12811 (53.5) | 11135 (46.5) | 12 | [9.3-15.0] | 56 | [41.5-67.9] | 2956 |
| Monogenic or congenital conditions |  |  |  |  |  |  |  |  |  |  |  |
| ADPKD | 119 (1.7) | 6993 (98.3) | 59 (49.6) | 60 (50.4) | 3302 (47.2) | 3691 (52.8) | 13 | [11.2-15.4] | 55 | [44.1-65.0] | 685 |
| ADTKD | ≤6 (NR*) | 190 (>97.0) | 0 (0.0) | 3 (100.0) | 80 (42.6) | 108 (57.4) | 17 | [15.7-17.2] | 54 | [43.3-63.9] | 24 |
| X-linked AS- Female | 46 (15.5) | 250 (84.5) | - | 46 (100.0) | - | 250 (100.0) | 11 | [8.7-14.6] | 46 | [33.3-59.3] | 13 |
| X-linked AS- Male | 53 (13.8) | 332 (86.2) | 53 (100.0) | - | 332 (100.0) | - | 12 | [10.0-14.9] | 43 | [29.4-56.4] | 27 |
| TBMN | 16 (9.9) | 146 (90.1) | ≤6 (NR) | 10 (>63.0) | 48 (32.9) | 98 (67.1) | 12 | [9.5-16.1] | 47 | [32.0-58.6] | 3 |
| ARPKD/NPHP | 71 (33.0) | 144 (67.0) | 40 (56.3) | 31 (43.7) | 64 (44.4) | 80 (55.6) | 12 | [9.2-14.5] | 39 | [26.5-54.8] | 18 |
| Cystinosis | 54 (37.5) | 90 (62.5) | 26 (48.1) | 28 (51.9) | 43 (47.8) | 47 (52.2) | 13 | [8.9-15.7] | 29 | [23.6-36.4] | 9 |
| Cystinuria | 28 (6.1) | 432 (93.9) | 19 (67.9) | 9 (32.1) | 222 (51.4) | 210 (48.6) | 11 | [7.2-14.5] | 49 | [34.4-62.1] | 13 |
| Hyperoxaluria | 25 (21.7) | 90 (78.3) | 12 (48.0) | 13 (52.0) | 59 (65.6) | 31 (34.4) | 11 | [9.0-13.3] | 36 | [26.4-52.1] | 9 |
| HNF1b Mutations | 31 (36.5) | 54 (63.5) | 19 (61.3) | 12 (38.7) | 28 (51.9) | 26 (48.1) | 9 | [6.8-14.2] | 39 | [23.7-51.1] | 1 |
| Renal Cancer Inherited | 10 (8.8) | 103 (91.2) | ≤6 (NR) | ≤6 (NR) | 39 (37.9) | 64 (62.1) | 10 | [8.4-12.5] | 54 | [35.4-60.8] | 1 |
| Tubulopathies | 76 (18.7) | 331 (81.3) | 54 (71.1) | 22 (28.9) | 155 (46.8) | 176 (53.2) | 12 | [8.1-15.9] | 40 | [29.5-56.0] | 12 |
| Tuberous Sclerosis Complex | 43 (17.8) | 199 (82.2) | 16 (37.2) | 27 (62.8) | 81 (40.7) | 118 (59.3) | 11 | [8.9-15.0] | 39 | [29.1-51.8] | 7 |
| Mostly non-monogenic or acquired conditions |  |  |  |  |  |  |  |  |  |  |  |
| aHUS | 89 (32.2) | 187 (67.8) | 43 (48.3) | 46 (51.7) | 85 (45.5) | 102 (54.5) | 10 | [7.0-13.9] | 42 | [32.7-56.4] | 17 |
| SSNS/MCD | 525 (31.8) | 1127 (68.2) | 329 (62.7) | 196 (37.3) | 638 (56.6) | 489 (43.4) | 12 | [9.3-14.2] | 47 | [31.5-63.6] | 53 |
| SRNS/FSGS | 256 (18.2) | 1154 (81.8) | 137 (53.5) | 119 (46.5) | 650 (56.3) | 504 (43.7) | 13 | [9.5-15.5] | 49 | [30.3-63.8] | 126 |
| INS-unspecified | 63 (7.4) | 792 (92.6) | 38 (60.3) | 25 (39.7) | 449 (56.7) | 343 (43.3) | 12 | [7.7-14.1] | 55 | [40.9-67.6] | 120 |
| IgA nephropathy | 40 (1.1) | 3756 (98.9) | 25 (62.5) | 15 (37.5) | 2610 (69.5) | 1146 (30.5) | 14 | [12.1-16.4] | 53 | [42.2-63.5] | 351 |
| Membranous Nephropathy | ≤6 (NR) | 2050 (>99.0) | ≤6 (NR) | ≤6 (NR) | 1358 (66.2) | 692 (33.8) | 14 | [13.3-15.1] | 66 | [55.5-74.6] | 384 |
| MGRS | 0 (0.0) | 144 (100.0) | 0 (0.0) | 0 (0.0) | 74 (51.4) | 70 (48.6) | - |  | 67 | [56.1-76.3] | 37 |
| MPGN/C3GN | 63 (6.8) | 869 (93.2) | 32 (50.8) | 31 (49.2) | 454 (52.2) | 415 (47.8) | 15 | [12.5-16.6] | 54 | [35.3-66.1] | 157 |
| Pregnancy | ≤6 (NR) | 680 (>99.0) | - | 1 (100.0) |  | 681 (100.0) | 16 | [16.1-16.1] | 37 | [33.1-42.0] | 10 |
| Retroperitoneal Fibrosis | 0 (0.0) | 111 (100.0) | 0 (0.0) |  | 72 (64.9) | 39 (35.1) | - |  | 67 | [58.5-74.2] | 31 |
| STEC HUS | 110 (65.9) | 57 (34.1) | 57 (51.8) | 53 (48.2) | 24 (42.1) | 33 (57.9) | 12 | [8.7-14.6] | 23 | [20.1-33.2] | 3 |
| ANCA associated vasculitis | 7 (0.4) | 1917 (99.6) | 0 (0.0) | 7 (100.0) | 1005 (52.4) | 912 (47.6) | 14 | [13.0-16.4] | 70 | [58.6-77.0] | 451 |
| Anti-GBM disease | ≤6 (NR) | 115 (>99.0) | 0 (0.0) | 1 (100.0) | 57 (49.6) | 58 (50.4) | 15 | [14.8-14.8] | 62 | [47.6-71.4] | 21 |
| Other Vasculitides | 200 (10.2) | 1757 (89.8) | 100 (50.0) | 100 (50.0) | 886 (50.4) | 871 (49.6) | 12 | [9.7-15.0] | 66 | [51.0-75.4] | 374 |

\*Prevalent patients on 25<sup>th</sup> July 2022. NR- Not reported; cells with fewer than 6 patients not reported due to risk of re-identification. Where a cell is not reported due to small numbers, corresponding cell values are rounded to the nearest 5. Individuals with two diagnoses are presented once for All RaDaR results, but subsequently included for each diagnosis. Row percentages are presented

The largest RDGs by patient number are ADPKD (n=7,112), Vasculitis (n=3,997), INS (n=3,917) and IgA nephropathy (n=3796). Conditions not presented due to low numbers are: Adenine phosphoribosyltransferase (APRT) deficiency (n=9), BK nephropathy (n=62), CKD due to Genetic factors in people of African Ancestry (CKD-AFRICA) (n=65), Calciphylaxis (n=59), Fabry Disease (n=47), Fibromuscular dysplasia (n=42), Mitochondrial renal disease (n=4) and Pure red cell aplasia (n=7).

Distribution of rare kidney diseases differed between patients currently ≤18 years old (paediatric) and >18 years old (adults). Predominant rare kidney diseases in adults were ADPKD (n=6993, 29%), Vasculitis (16%; ANCA associated vasculitis n=1917, Anti-GBM disease n=115, Other Vasculitides n=1757) and IgA nephropathy (n= 3756, 16%). In children the largest RDGs were Idiopathic Nephrotic Syndrome (44%; SSNS/MCD n= 525, SRNS/FSGS n=256, INS-unspecified n= 63), Vasculitis (11%; ANCA associated vasculitis n=7, Anti-GBM disease ≤6, Other Vasculitides n=200) and Alport Syndrome (6%; X-linked Males n=53 X-linked Females n=46 TBMN n=16). The most common rare kidney diseases at time of diagnosis in adults and children were the same.

Males were over-represented among paediatric patients with Cystinuria (68%), HNF1b mutations (61%), INS (60%) and Tubulopathies (71%) (due to male predominant Lowe Syndrome); among adult patients with Membranous Nephropathy (66%), Retroperitoneal Fibrosis (65%) and Primary Hyperoxaluria (66%); and among both children and adults with IgA nephropathy (children 63%, adults 70%). Minimum UK point prevalence and maximum area density estimates per 100,000 population for each RDG are presented in Table 2.

**Table 2:**
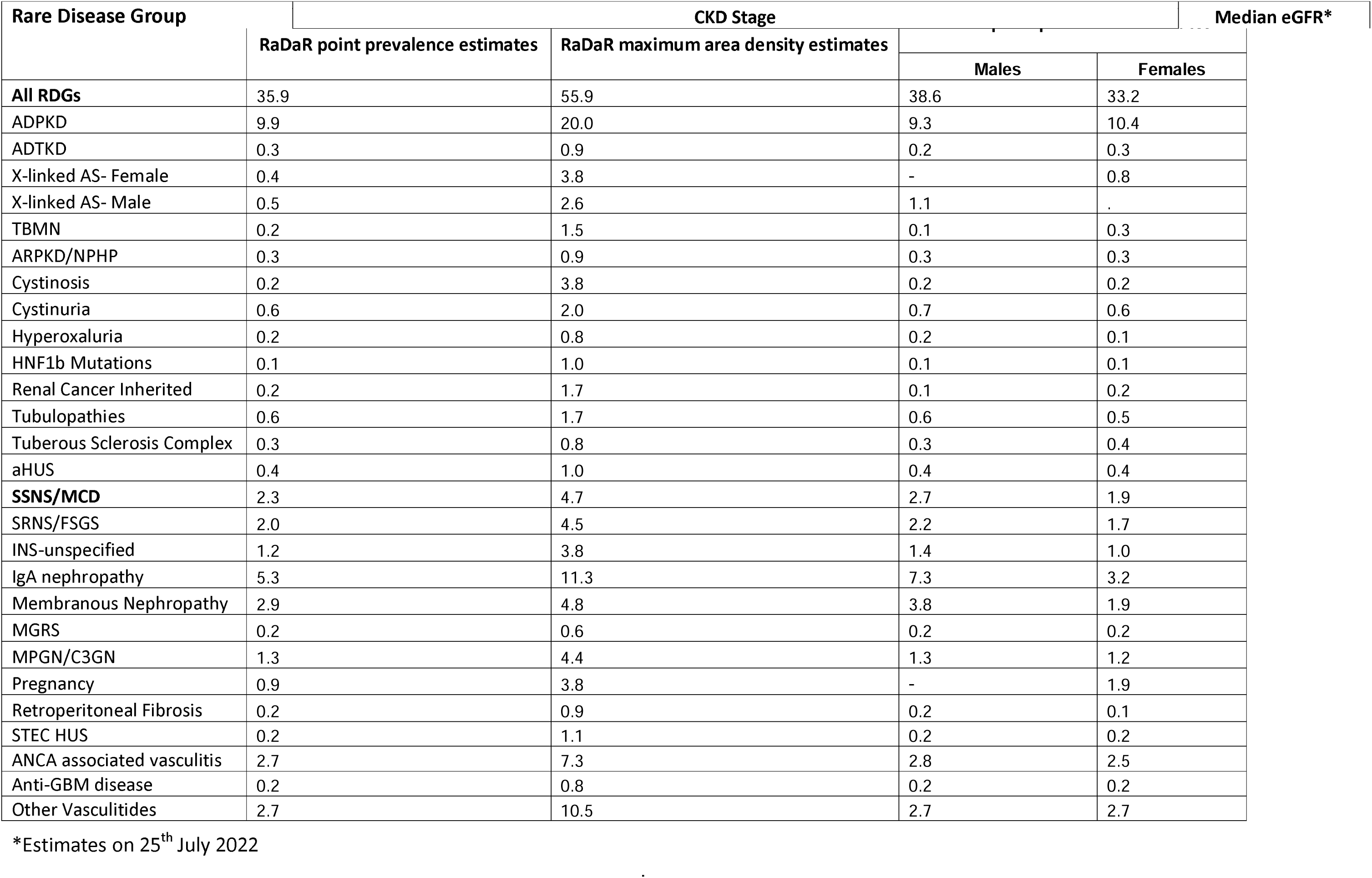
RaDaR point prevalence rates and maximum area density estimates per 100,000 population*, stratified by RDG and sex.

Ethnic distribution and socioeconomic status for all RaDaR patients, stratified by RDG are presented in Figure 2. Excluding patients with missing data, 87% of RaDaR participants are White, 1% Mixed, 8% Asian, 3% Black and 1% Other ethnicities. The SSNS/MCD, SRNS/FSGS, IgA nephropathy, Tubulopathies, Cystinosis and Primary Hyperoxaluria RDGs all had a significantly larger proportion of patients from Asian ethnic backgrounds than the total RaDaR population. This was particularly marked in the Cystinosis and Primary Hyperoxaluria RDGs where the proportion of patients from Asian backgrounds was higher than the overall RaDaR proportion by 17% (95% CI: 8%-25%) and 27% (95% CI: 17%-37%) respectively. Similar differences were observed when stratifying by paediatric and adult populations (Supplementary Figures 1-2). Individuals recruited to the pregnancy RDG were also more likely to be from Asian or Black backgrounds, 8% (95% CI: 5%-12%) and 9% (95% CI: 6%-12%) increase respectively. Children recruited to RaDaR from all 4 UK nations were more likely to be from Asian ethnic backgrounds when compared to adults recruited to RaDaR (17% vs 8%, p<0.0001, Supplementary Table 5), and also when English children recruited to RaDaR were compared to children in the general English population (18% vs 10%, p-value <0.0001, Supplementary Table 6) 38% of patients recruited to RaDaR had monogenic disorders (disorders usually caused by inheritance of one or two pathogenic variants in a single gene, or a *de novo* mutation). Stratifying by ethnicity, most monogenic disorders were diagnosed in White patients (90.3%) (Supplementary Table 7). Adults recruited to RaDaR were more likely to be diagnosed with monogenic disorders than children (39% vs 30%, p-value <0.0001, Supplementary Table 8). Only adults from White, Mixed and Black backgrounds were more likely to be diagnosed with monogenic disorders compared to children; children from Asian or Other ethnic backgrounds are as likely to have a diagnosis of a monogenic disorder as adults (24% vs 23%, p-value 0.7 and 33% vs 32%, p-value 0.9 respectively).

**Figure 2:**
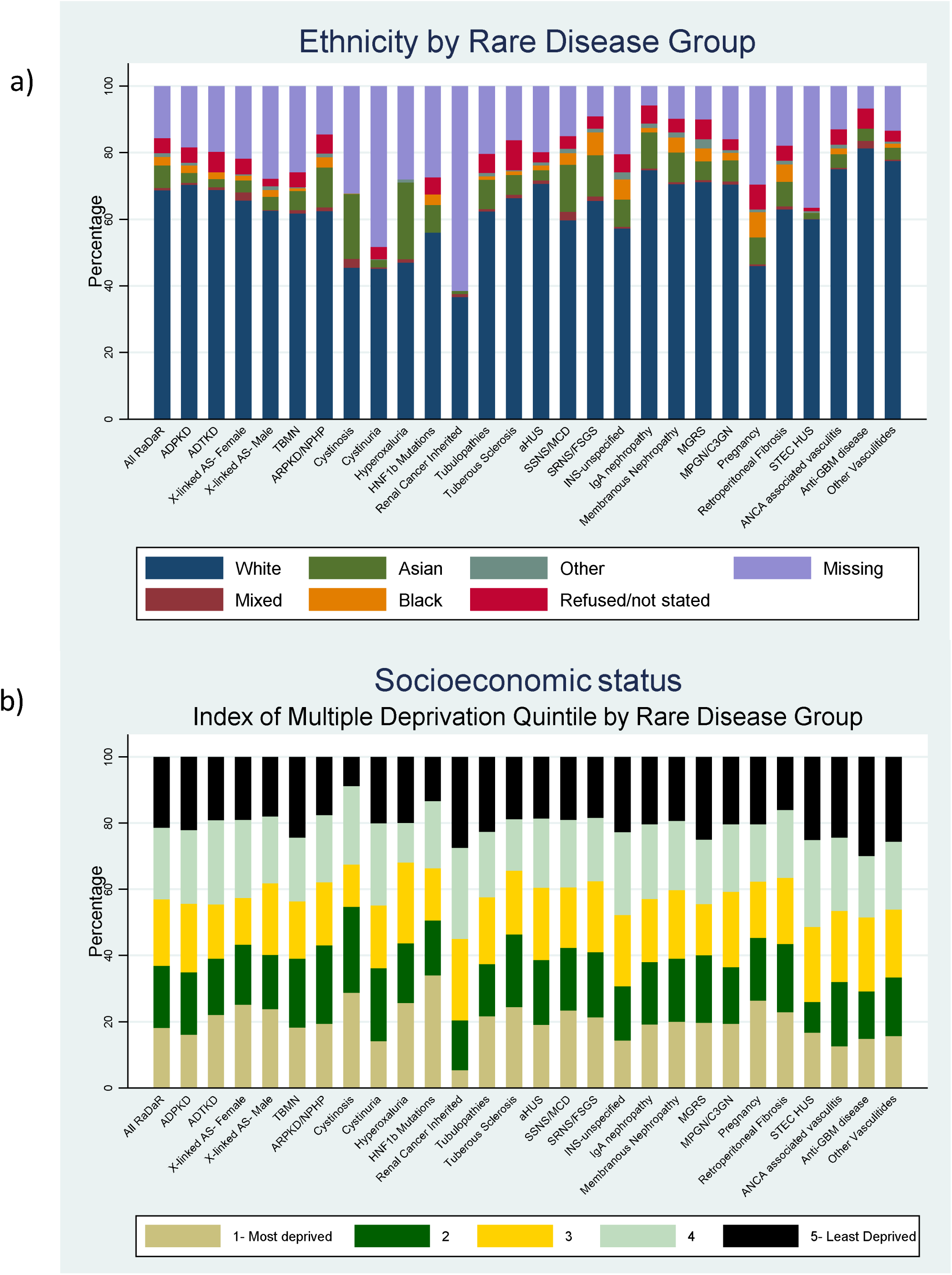
RaDaR patient a) Ethnicity and b) Socioeconomic status, stratified by Rare Disease Gro

Socioeconomic deprivation varied by RDG; patients diagnosed with Cystinosis, Primary Hyperoxaluria, SSNS/MCD, SRNS/FSGS, IgA nephropathy, Membranous Nephropathy and Pregnancy were more likely to be in the most deprived IMD Quintile compared to the overall RaDaR population. There were similar differences in both adult and paediatric patients (Supplementary Figures 3-4). Individuals with non-monogenic disorders were more likely to be in the most deprived IMD Quintile compared to those with monogenic disorders, but this association was attenuated when stratifying by ethnicity (Supplementary Table 9). Individuals with autosomal dominant conditions were less likely to be in the most deprived quintile compared to those with autosomal recessive, X-linked or non-monogenic disorders (16% vs 20% vs 19% respectively, p-value <0.0001, Supplementary Table 10).

More paediatric patients were in the most deprived IMD Quintile when compared to adults recruited to RaDaR (30% vs 17%, Supplementary Table 5), and when compared to children in the general English population (Supplementary Table 6). Paediatric patients of White, Asian, and Other ethnicities were all more likely to be in the most deprived IMD quintile compared to adults (25% vs 16% p<0.0001, 54% vs 31% p<0.0001, 50% vs 25% p=0.02, respectively, Supplementary Table 11). Children with monogenic and non-monogenic disorders, and those with monogenic disorders with all modes of inheritance were more likely to be in the most deprived quintile compared to adults (Supplementary Tables 10 and 12).

### Renal Function of the RaDaR patient population

Many patients in RaDaR had reached KF (CKD stage G5 or KRT) (39%) (Table 3). This proportion varied by RDG; only 2% of patients with Cystinuria had reached KF compared to 73% of male patients with AS, or 70% of patients with Cystinosis. Most paediatric patients had eGFR results >60ml/min/1.73m^2^ (71% CKD stages G1-G2 vs 32% adult patients).

**Table 3:**
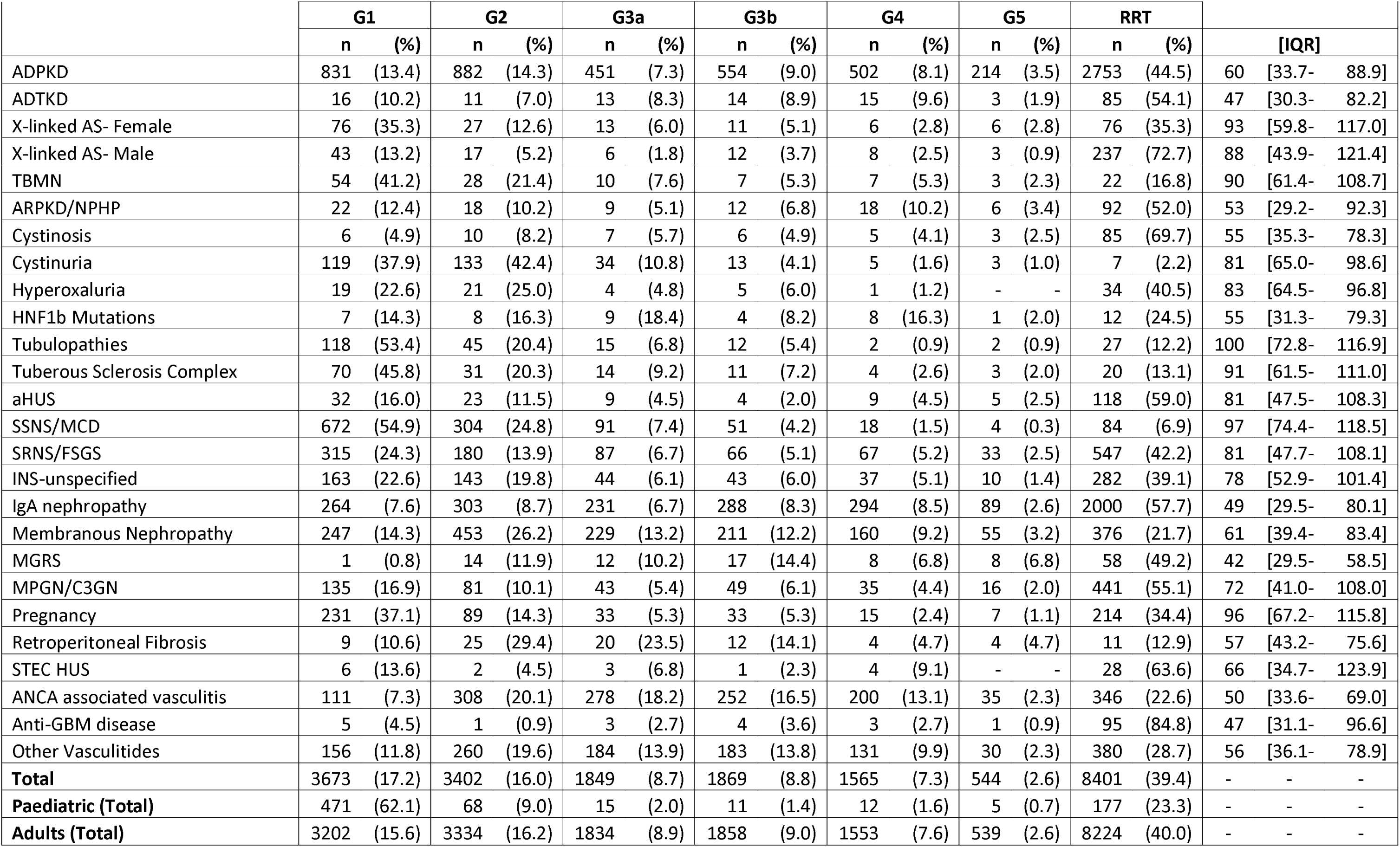
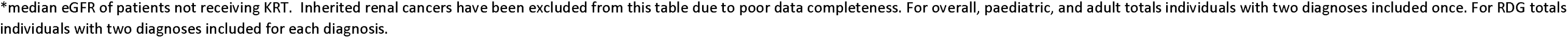
CKD stage and Median eGFR of RaDaR patients on 1^st^ January 2022.

|  | G1 |  | G2 |  | G3a |  | G3b |  | G4 |  | G5 |  | RRT |  |  |  |  |
| --- | --- | --- | --- | --- | --- | --- | --- | --- | --- | --- | --- | --- | --- | --- | --- | --- | --- |
|  | n | (%) | n | (%) | n | (%) | n | (%) | n | (%) | n | (%) | n | (%) | [IQR] |  |  |
| ADPKD | 831 | (13.4) | 882 | (14.3) | 451 | (7.3) | 554 | (9.0) | 502 | (8.1) | 214 | (3.5) | 2753 | (44.5) | 60 | [33.7- | 88.9] |
| ADTKD | 16 | (10.2) | 11 | (7.0) | 13 | (8.3) | 14 | (8.9) | 15 | (9.6) | 3 | (1.9) | 85 | (54.1) | 47 | [30.3- | 82.2] |
| X-linked AS- Female | 76 | (35.3) | 27 | (12.6) | 13 | (6.0) | 11 | (5.1) | 6 | (2.8) | 6 | (2.8) | 76 | (35.3) | 93 | [59.8- | 117.0] |
| X-linked AS- Male | 43 | (13.2) | 17 | (5.2) | 6 | (1.8) | 12 | (3.7) | 8 | (2.5) | 3 | (0.9) | 237 | (72.7) | 88 | [43.9- | 121.4] |
| TBMN | 54 | (41.2) | 28 | (21.4) | 10 | (7.6) | 7 | (5.3) | 7 | (5.3) | 3 | (2.3) | 22 | (16.8) | 90 | [61.4- | 108.7] |
| ARPKD/NPHP | 22 | (12.4) | 18 | (10.2) | 9 | (5.1) | 12 | (6.8) | 18 | (10.2) | 6 | (3.4) | 92 | (52.0) | 53 | [29.2- | 92.3] |
| Cystinosis | 6 | (4.9) | 10 | (8.2) | 7 | (5.7) | 6 | (4.9) | 5 | (4.1) | 3 | (2.5) | 85 | (69.7) | 55 | [35.3- | 78.3] |
| Cystinuria | 119 | (37.9) | 133 | (42.4) | 34 | (10.8) | 13 | (4.1) | 5 | (1.6) | 3 | (1.0) | 7 | (2.2) | 81 | [65.0- | 98.6] |
| Hyperoxaluria | 19 | (22.6) | 21 | (25.0) | 4 | (4.8) | 5 | (6.0) | 1 | (1.2) | - | - | 34 | (40.5) | 83 | [64.5- | 96.8] |
| HNF1b Mutations | 7 | (14.3) | 8 | (16.3) | 9 | (18.4) | 4 | (8.2) | 8 | (16.3) | 1 | (2.0) | 12 | (24.5) | 55 | [31.3- | 79.3] |
| Tubulopathies | 118 | (53.4) | 45 | (20.4) | 15 | (6.8) | 12 | (5.4) | 2 | (0.9) | 2 | (0.9) | 27 | (12.2) | 100 | [72.8- | 116.9] |
| Tuberous Sclerosis Complex | 70 | (45.8) | 31 | (20.3) | 14 | (9.2) | 11 | (7.2) | 4 | (2.6) | 3 | (2.0) | 20 | (13.1) | 91 | [61.5- | 111.0] |
| aHUS | 32 | (16.0) | 23 | (11.5) | 9 | (4.5) | 4 | (2.0) | 9 | (4.5) | 5 | (2.5) | 118 | (59.0) | 81 | [47.5- | 108.3] |
| SSNS/MCD | 672 | (54.9) | 304 | (24.8) | 91 | (7.4) | 51 | (4.2) | 18 | (1.5) | 4 | (0.3) | 84 | (6.9) | 97 | [74.4- | 118.5] |
| SRNS/FSGS | 315 | (24.3) | 180 | (13.9) | 87 | (6.7) | 66 | (5.1) | 67 | (5.2) | 33 | (2.5) | 547 | (42.2) | 81 | [47.7- | 108.1] |
| INS-unspecified | 163 | (22.6) | 143 | (19.8) | 44 | (6.1) | 43 | (6.0) | 37 | (5.1) | 10 | (1.4) | 282 | (39.1) | 78 | [52.9- | 101.4] |
| IgA nephropathy | 264 | (7.6) | 303 | (8.7) | 231 | (6.7) | 288 | (8.3) | 294 | (8.5) | 89 | (2.6) | 2000 | (57.7) | 49 | [29.5- | 80.1] |
| Membranous Nephropathy | 247 | (14.3) | 453 | (26.2) | 229 | (13.2) | 211 | (12.2) | 160 | (9.2) | 55 | (3.2) | 376 | (21.7) | 61 | [39.4- | 83.4] |
| MGRS | 1 | (0.8) | 14 | (11.9) | 12 | (10.2) | 17 | (14.4) | 8 | (6.8) | 8 | (6.8) | 58 | (49.2) | 42 | [29.5- | 58.5] |
| MPGN/C3GN | 135 | (16.9) | 81 | (10.1) | 43 | (5.4) | 49 | (6.1) | 35 | (4.4) | 16 | (2.0) | 441 | (55.1) | 72 | [41.0- | 108.0] |
| Pregnancy | 231 | (37.1) | 89 | (14.3) | 33 | (5.3) | 33 | (5.3) | 15 | (2.4) | 7 | (1.1) | 214 | (34.4) | 96 | [67.2- | 115.8] |
| Retroperitoneal Fibrosis | 9 | (10.6) | 25 | (29.4) | 20 | (23.5) | 12 | (14.1) | 4 | (4.7) | 4 | (4.7) | 11 | (12.9) | 57 | [43.2- | 75.6] |
| STEC HUS | 6 | (13.6) | 2 | (4.5) | 3 | (6.8) | 1 | (2.3) | 4 | (9.1) | - | - | 28 | (63.6) | 66 | [34.7- | 123.9] |
| ANCA associated vasculitis | 111 | (7.3) | 308 | (20.1) | 278 | (18.2) | 252 | (16.5) | 200 | (13.1) | 35 | (2.3) | 346 | (22.6) | 50 | [33.6- | 69.0] |
| Anti-GBM disease | 5 | (4.5) | 1 | (0.9) | 3 | (2.7) | 4 | (3.6) | 3 | (2.7) | 1 | (0.9) | 95 | (84.8) | 47 | [31.1- | 96.6] |
| Other Vasculitides | 156 | (11.8) | 260 | (19.6) | 184 | (13.9) | 183 | (13.8) | 131 | (9.9) | 30 | (2.3) | 380 | (28.7) | 56 | [36.1- | 78.9] |
| **Total** | 3673 | (17.2) | 3402 | (16.0) | 1849 | (8.7) | 1869 | (8.8) | 1565 | (7.3) | 544 | (2.6) | 8401 | (39.4) | - | - | - |
| **Paediatric (Total)** | 471 | (62.1) | 68 | (9.0) | 15 | (2.0) | 11 | (1.4) | 12 | (1.6) | 5 | (0.7) | 177 | (23.3) | - | - | - |
| **Adults (Total)** | 3202 | (15.6) | 3334 | (16.2) | 1834 | (8.9) | 1858 | (9.0) | 1553 | (7.6) | 539 | (2.6) | 8224 | (40.0) | - | - | - |

\*median eGFR of patients not receiving KRT. Inherited renal cancers have been excluded from this table due to poor data completeness. For overall, paediatric, and adult totals individuals with two diagnoses included once. For RDG totals individuals with two diagnoses included for each diagnosis.

### Recruitment to RaDaR

Geographic distribution of recruitment to RaDaR across the UK is shown in Figure 3. Comparison of RaDaR with UKRR rare disease KRT populations (Figure 4 and Supplementary Table 13) demonstrated similar distributions in both populations, and testing for a difference found negligible evidence (Cramer’s V = 0.07)

**Figure 3:**
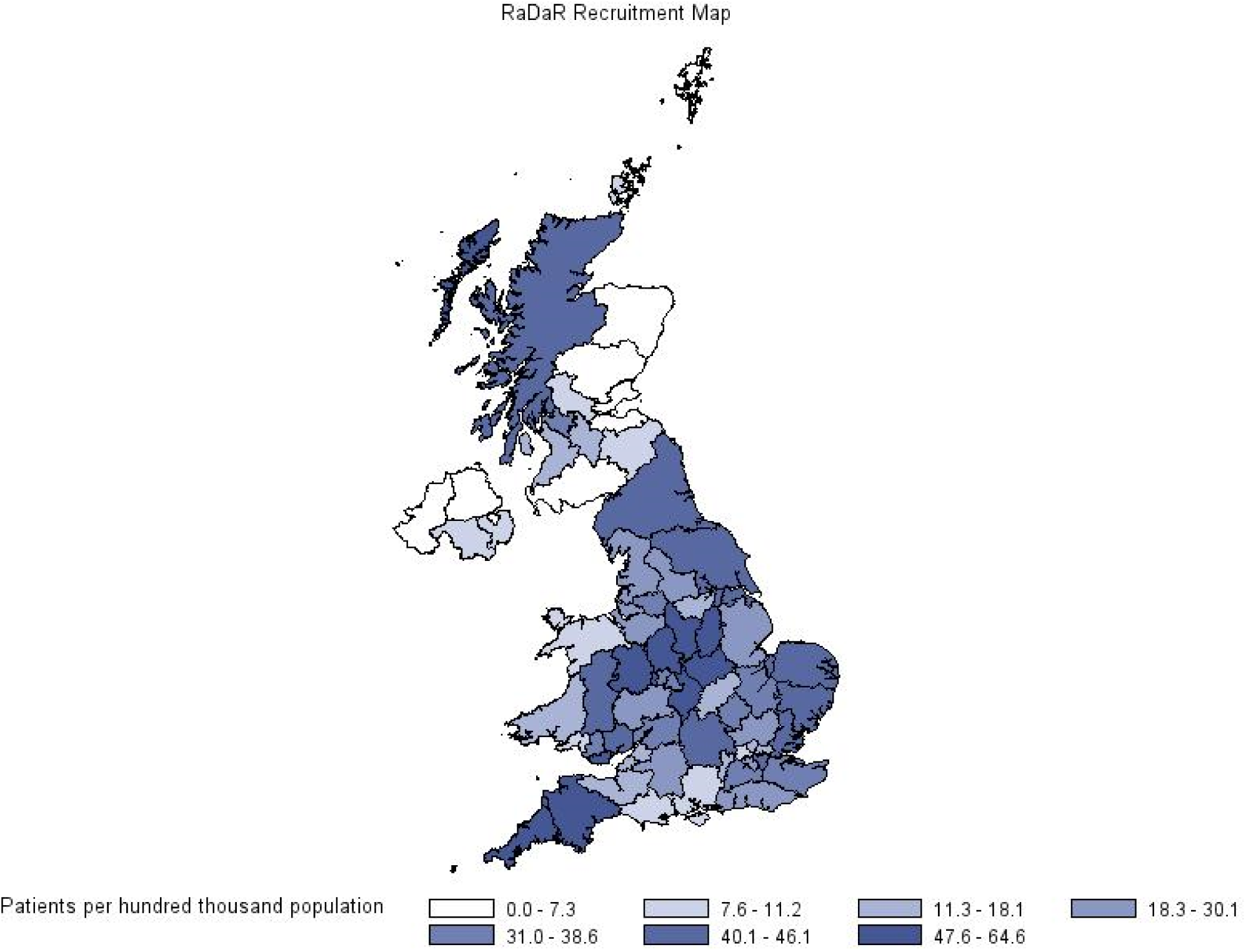
Distribution of recruitment to RaDaR across the United Kingdom

**Figure 4:**
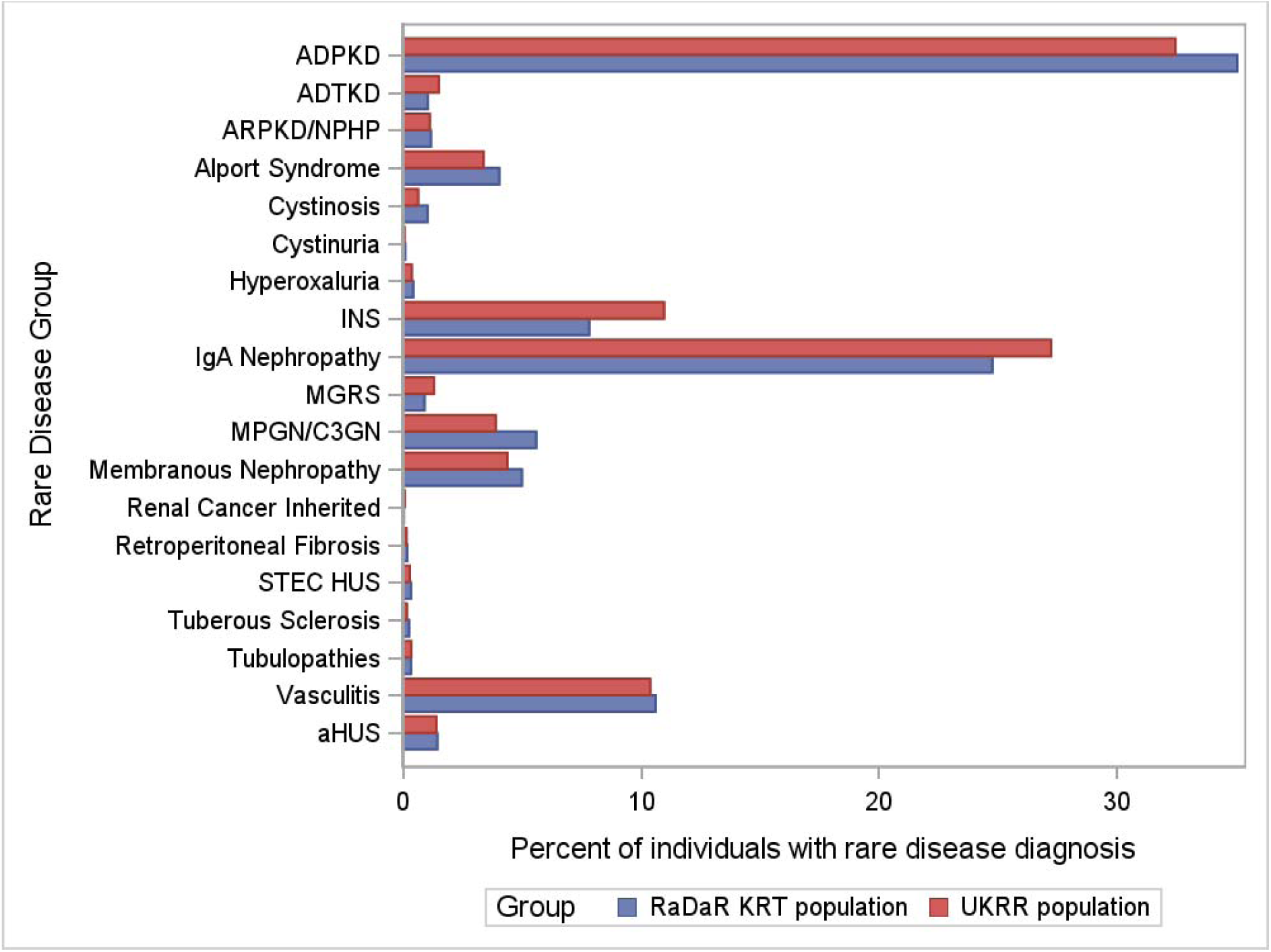
Proportion of rare disease diagnoses for each disorder within UKRR and RaDaR KRT recipients

Overall, patients recruited to RaDaR with KF were less likely to be Asian (7% vs 9%, p<0.0001) when compared with unrecruited patients in the UKRR dataset (Table 4). However, there was no significant difference between the ethnic distribution of recruited vs. unrecruited patients in the following RDGs: ARPKD (Chi^2^ p=0.41), AS (p=0.76), atypical HUS (p=0.55), Cystinosis (p=0.71), Primary Hyperoxaluria (p=0.49), MPGN/C3GN (p=0.15), STEC-HUS (p=0.62), and Membranous Nephropathy (p=0.44). UKRR patients in the least deprived quintile were more likely to be recruited to RaDaR than those in the most deprived quintile (21% vs 17%, p<0.0001).

**Table 4:**
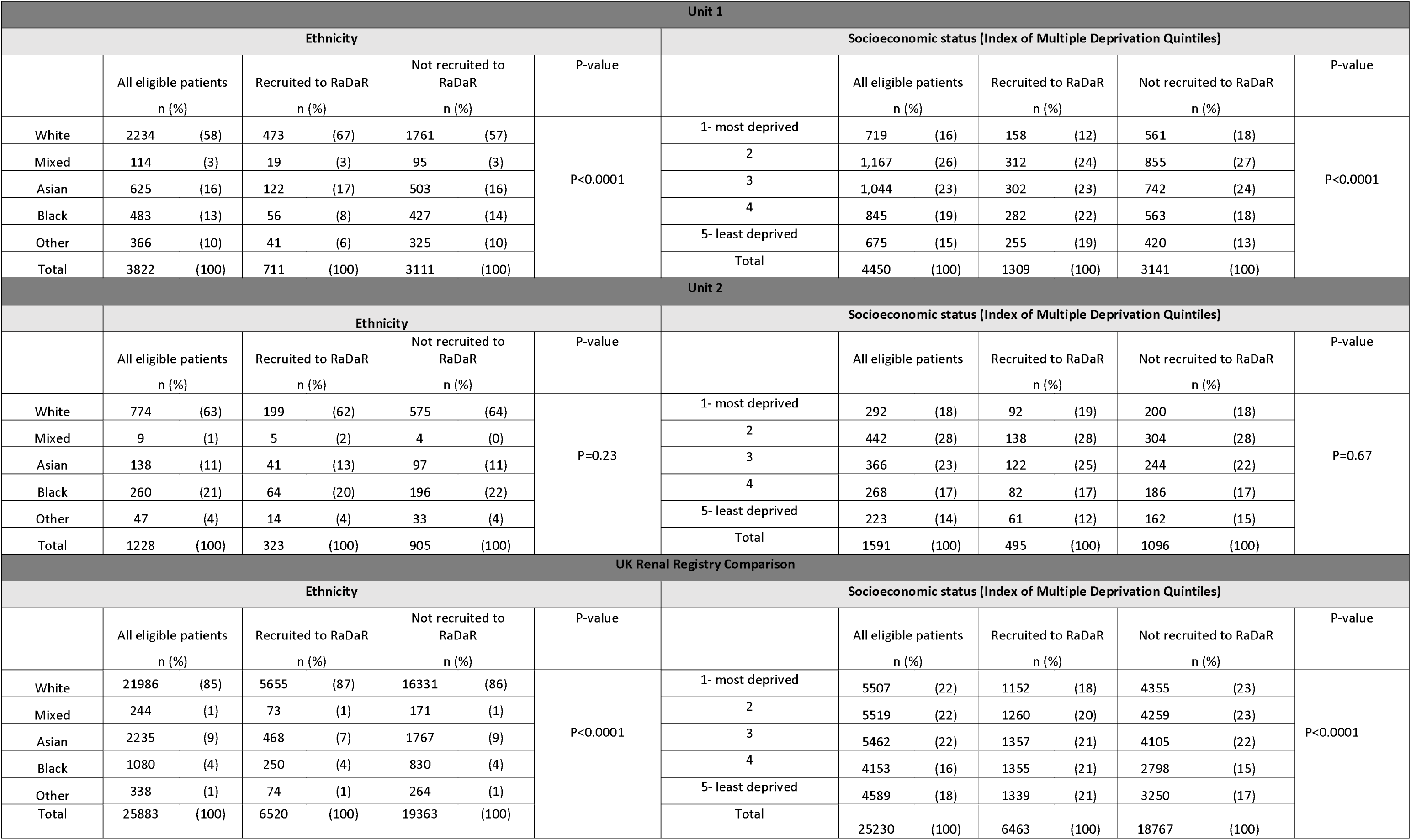
Ethnicity and Socioeconomic status comparisons between RaDaR recruited vs unrecruited patients with rare kidney diagnoses at 2 UK renal units and between recruited patients with Kidney Failure and UKRR patients with rare kidney diseases.

Results comparing recruitment to RaDaR at two large renal centres were conflicting; comparing recruited and non-recruited patients at each centre, and excluding patients with ethnicity not recorded, there were no differences in ethnicity observed in one centre (recruited vs non recruited, White 62% vs 64%, Mixed 2% vs 0%, Asian 13% vs 11%, Black 20% vs 22%, Other ethnicity 4% vs 4%, p<0.23), and evidence of over-recruitment of White (67% vs 57%, p<0.0001) and under-recruitment of Black patients (8% vs 14%, p<0.0001) in the other centre. Similarly, there was no evidence of difference in social deprivation in one centre (recruited vs non recruited, IMD Quintile 1-most deprived: 19% vs 18%, Quintile 2: 28% vs 28%, Quintile 3: 25% vs 22%, Quintile 4: 17% vs 17%, Quintile 5-least deprived: 12% vs 15% , p<0.67), whereas there was evidence of over-recruitment of patients in the least deprived quintile (19% vs 13%, p<0.0001) in the other.

Prevalent English patients with a diagnosis of ADPKD were compared with the 2011 English Census, adjusted for age (Supplementary Figure 3). Deviation from the ethnic distribution of the English population varied by age-group. More patients aged <40 years were White than the general population (0-17 years 83% vs 79% p=0.006, 18-29 years 92% vs 81% p=0.001, 30-39 years 92% vs 81% p<0.0001). However, patients aged 60-69 years were more likely to be Black than the general population (3% vs 1%, p<0.0001). There was no difference in ethnic distribution for individuals more than 80 years old (p=0.39).

## Discussion

We have presented cross-sectional analyses for 1,934 (7%) paediatric and 23,946 (93%) adult patients with rare kidney diseases enrolled into RaDaR. To our knowledge, this is the largest epidemiological description of rare kidney diseases worldwide.

RaDaR is not a population-based registry, and therefore cannot offer precise incidence or prevalence data on individual rare kidney disorders; however, RaDaR patient numbers have allowed us to provide minimum point prevalence UK estimates for 21 RDGs, in some cases for the first time. Additionally, patient numbers and demographics may be useful in assessing feasibility of studies or clinical trials in individual rare kidney diseases. Comparison of RaDaR KRT recipients with UK recipients of KRT with a rare disease recorded in UKRR indicated that a similar proportion (approximately 1:2-1:3) of all eligible rare disease patients with KF were enrolled in RaDaR, indicating that in this group RaDaR allows comparison of prevalence of the relevant diseases without detectable bias according to disease. Similar analyses could not be done in the non-KRT population because there is no comprehensive registry in this group to compare with.

After comparison with multiple data sources, we found that while there was likely to be imperfect representation of all socio-economic and ethnic groups in RaDaR, these biases do not appear to be of a magnitude likely to distort inferences about epidemiology or natural history of RaDaR diseases. Whilst patients with KF in RaDaR are more likely to be White than eligible patients in the UKRR, comparison of the English RaDaR ADPKD cohort with the English census and comparison with patients two large renal units found no consistent ethnic recruitment bias to RaDaR. For seven RDGs (ARPKD, Alport syndrome, atypical HUS, STEC-HUS, cystinosis, primary hyperoxaluria, MPGN/C3GN and membranous nephropathy) patients with KF recruited to RaDaR closely matched the ethnic distribution of patients in the UKRR. Patients recruited to RaDaR were more likely to be from the least deprived quintile compared to the UKRR, whereas there was no evidence of over-recruitment of patients in the least deprived quintile within a large renal unit. Future work will include investigating these differences to identify potential inequity and to target future recruitment strategies.

Despite evidence that patients of white ethnicities may be over-represented in the RaDaR KF population, we found patients with cystinosis and primary hyperoxaluria were less likely to be White, and more likely to be from Asian backgrounds compared to the overall ethnic distribution of RaDaR. These differences were also present when stratifying by paediatric and adult RaDaR patients, although paediatric data should be interpreted with caution due to small patient numbers. This ethnic predisposition has been previously reported in cystinosis, with a high birth frequency rate (1:3,600) reported in Pakistani ethnic groups in the West Midlands^17^. However, to our knowledge it has not previously been reported in primary hyperoxaluria (PH). Previous population analyses have suggested PH is 3 times more prevalent amongst European Americans than African Americans^18^, and that certain PH1 gene variants have a strong association with people from Spanish or North African backgrounds^19^. Whilst a possible mutational hotspot in PH3 gene *HOGA1* has been identified in the Chinese population^20^, none of the patients in the RaDaR PH cohort were from a Chinese background. As for any autosomal recessive diseases, the frequency of consanguinity in the community may impact on the incidence of cystinosis and PH.

Paediatric patients were more likely to be from Asian backgrounds compared to adults. This is likely due to the higher proportion of INS (SSNS, SRNS and INS-unspecified) in the paediatric group (44% children vs. 13% adults), conditions that have been reported to affect South Asians up to five times more frequently than Europeans^21,22^.

More paediatric patients were in the lowest IMD quintile compared to adults. In the UK, children are more likely to live in more deprived areas compared to adults, and people of Asian and Black ethnicity are more likely to live in areas with the worst levels of social deprivation than those of White ethnicity^23^. However, the proportion of English children recruited to RaDaR living in the most deprived IMD quintile exceeded that of children in the general English population. Differences in ethnicity and the proportion of monogenic and non-monogenic conditions between the paediatric and adult RaDaR populations do not completely explain this disparity; children from White, Asian and other ethnicities were all more likely to live in more socially deprived areas than adults, as were children diagnosed with both monogenic and non-monogenic disorders. Paediatric patients may be more intensively recruited to RaDaR from centres in areas of worst deprivation, either due to clinician interest or a higher population prevalence of certain rare kidney diseases in those areas. Rare diseases are associated with a high economic burden for patients, especially for families with children^24^. Children from socioeconomically deprived backgrounds experience poorer health outcomes^25,26^, and there is evidence of reduced access to pre-emptive kidney transplantation in UK paediatric kidney patients from more deprived areas^27^. These findings therefore highlight that children with rare kidney diseases recruited to RaDaR are a potentially highly vulnerable group; further investigation is needed to determine whether they experience different outcomes.

### Limitations

RaDaR is a UK registry and is representative of the mainly White UK population and may not be generalisable to other ethnicities. Survivor bias may have had an impact on the enrolment of individuals with diagnoses made before RaDaR started recruiting patients with that condition.

Some RaDaR diagnoses are poorly captured by ERA-EDTA PRD codes, which limited comparison to UKRR data. Entry of rare disease diagnoses into renal IT systems is user dependent and may vary between renal units used for comparisons.

Although bias could be introduced owing to the variation in recruitment between centres across the UK, and therefore by variation in their catchment population, we sought to minimise effects of this bias by comparing ethnicity and SES of each RDG to the overall RaDaR breakdown. Caution must still be exercised where clinicians with particular interest in a certain RDG recruit more intensively into that one RDG compared to others.

In summary, to our knowledge, RaDaR is the largest registry of rare kidney diseases worldwide and provides numerous opportunities to study rare kidney diseases, including identification of potential participants in clinical trials.

## Supporting information

Supplementary appendix 1 Radar consortium

Supplementary appendix 2 Inclusion-Exclusion criteria

## Data Availability

The minimal underlying dataset cannot be included as it contains confidential, potentially identifying and sensitive patient information. The UK Kidney Association(UKKA) is the controller of the data and restricts sharing of individual level data if there is no direct relationship between the UKKA and the recipient. The data underlying these results are available for researchers
who meet the criteria for access to confidential
data.

