## Supplementary Materials Tables and Figures for "The National Registry of Rare Kidney Diseases (RaDaR): Description, recruitment, and cross-sectional analyses of 25,880 adults and children with rare kidney diseases in the UK"

**Supplementary Methods**

**Supplementary Table 1:** Missing data analysis for individuals missing date of diagnosis

**Supplementary Table 2:** Missing data analysis for individuals missing ethnicity

**Supplementary Table 3:** Missing data analysis for individuals missing IMD quintile data

**Supplementary Table 4:** ERA-EDTA codes and keyword search terms for each Rare Disease Group

**Supplementary Table 5:** Ethnicity and IMD Quintile, stratified by current age

**Supplementary Table 6:** Ethnicity and IMD Quintile distribution of children (aged ≤18 years) in England compared to the children in RaDaR

**Supplementary Table 7:** Monogenic and Non monogenic disorders, stratified by ethnicity

**Supplementary Table 8:** Monogenic and Non monogenic disorders, stratified by age category and ethnicity

**Supplementary Table 9:** Monogenic and Non monogenic disorders, stratified by Ethnicity and IMD Quintile

**Supplementary Table 10:** Socioeconomic status (IMD Quintile) of RaDaR participants, stratified by mode of inheritance and age category

**Supplementary Table 11:** IMD Quintile, stratified by current age and Ethnicity

**Supplementary Table 12:** Socioeconomic status (IMD Quintile), stratified by monogenic and non monogenic disorders and age category*

**Supplementary Table 13:** Number and percentage of rare disease diagnoses for each disorder within UKRR and RaDaR KRT recipients

**Supplementary Figure 1:** Comparison of ethnicity in each Rare Disease Group to total ethnicity breakdown of RaDaR

**Supplementary Figure 2:** Comparison of ethnicity in each Rare Disease Group to total ethnicity breakdown of RaDaR, stratified by age group

**Supplementary Figure 3:** Comparison of IMD Quintile in each Rare Disease Group to total IMD Quintile distribution of RaDaR

**Supplementary Figure 4:** Comparison of ethnicity in each Rare Disease Group to total ethnicity breakdown of RaDaR, stratified by age group

**Supplementary Figure 5:** Ethnicity of English RaDaR patients with ADPKD compared to English census

**Supplementary Methods**

To assess whether there has been ethnic or socioeconomic status (SES) recruitment bias to RaDaR, three methods were used. Firstly, ethnicity and SES of all prevalent RaDaR patients who had reached kidney failure were compared to patients with a rare kidney diagnosis in the UKRR. All UKRR patients from renal units participating in RaDaR, alive after their renal unit began recruiting to RaDaR, and not already recruited to RaDaR were then identified using the RaDaR list of PRD codes. To ensure an accurate comparison, only RDGs where RaDaR diagnoses were well captured by the UKRR, and where there was unlikely to be a difference between the diagnosis given at start of RRT and a patient’s RaDaR diagnosis were included. These RDGs were determined by ascertaining 1) RDGs where >60% of recruited patients with ESKD were identified using the overall RaDaR PRD code list in the UKRR dataset 2) RDGs where <30% of patients had a diagnosis date after commencement of RRT.

Secondly, patients recruited to RaDaR from two large UK renal centres were compared with all unrecruited patients with a RaDaR eligible diagnosis at those centres, alive after those centres began participating in RaDaR (personal communication with KW). Eligible patients were identified from renal IT systems using PRD codes or diagnosis keywords (Supplementary Table 4).

Lastly, the age-stratified ethnicity distribution of England according to the 2011 UK census was compared to the ethnicity of prevalent English RaDaR patients with ADPKD. As a hereditary condition driven by recent or de-novo mutations, ADPKD is likely to affect ethnic groups equally^1,2^; ethnic distribution is therefore expected to reflect that reported by the UK census. Persistent deviation from the ethnic distribution reported by the census in this RDG might therefore suggest bias in recruitment to RaDaR.

**Supplementary Table 1: Missing data analysis for individuals missing date of diagnosis**

|  | **Not missing** | | **Missing** | | | **P-value*** |
| --- | --- | --- | --- | --- | --- | --- |
|  | **n** | **(%)** | | **n** | **(%)** |  |
| **Cohort** | | | | | | |
| ADPKD | 5008 | (24.0) | | 2104 | (40.9) | <0.0001 |
| ADTKD | 155 | (0.7) | | 36 | (0.7) |  |
| X-linked AS- Female | 241 | (1.2) | | 55 | (1.1) |  |
| X-linked AS- Male | 306 | (1.5) | | 79 | (1.5) |  |
| TBMN | 154 | (0.7) | | 8 | (0.2) |  |
| ARPKD/NPHP | 167 | (0.8) | | 48 | (0.9) |  |
| Cystinosis | 129 | (0.6) | | 15 | (0.3) |  |
| Cystinuria | 409 | (2.0) | | 51 | (1.0) |  |
| Hyperoxaluria | 96 | (0.5) | | 19 | (0.4) |  |
| HNF1b Mutations | 68 | (0.3) | | 17 | (0.3) |  |
| Renal Cancer Inherited | 112 | (0.5) | | 1 | (0.0) |  |
| Tubulopathies | 305 | (1.5) | | 102 | (2.0) |  |
| Tuberous Sclerosis Complex | 201 | (1.0) | | 41 | (0.8) |  |
| aHUS | 221 | (1.1) | | 55 | (1.1) |  |
| SSNS/MCD | 1621 | (7.8) | | 31 | (0.6) |  |
| SRNS/FSGS | 1370 | (6.6) | | 40 | (0.8) |  |
| INS- unspecified | 369 | (1.8) | | 486 | (9.4) |  |
| IgA nephropathy | 3398 | (16.3) | | 398 | (7.7) |  |
| Membranous Nephropathy | 1670 | (8.0) | | 385 | (7.5) |  |
| MGRS | 122 | (0.6) | | 22 | (0.4) |  |
| MPGN/C3GN | 739 | (3.5) | | 193 | (3.7) |  |
| Pregnancy | 467 | (2.2) | | 215 | (4.2) |  |
| Retroperitoneal Fibrosis | 87 | (0.4) | | 24 | (0.5) |  |
| STEC HUS | 151 | (0.7) | | 16 | (0.3) |  |
| ANCA associated vasculitis | 1920 | (9.2) | | 4 | (0.1) |  |
| Anti-GBM disease | 116 | (0.6) | | 0 | (0.0) |  |
| Other Vasculitides | 1252 | (6.0) | | 705 | (13.7) |  |
| **Ethnicity** | | | | | | |
| White | 14311 | (68.9) | | 3977 | (77.8) | <0.0001 |
| Mixed | 166 | (0.8) | | 32 | (0.6) |  |
| Asian | 1491 | (7.2) | | 235 | (4.6) |  |
| Black | 505 | (2.4) | | 113 | (2.2) |  |
| Other | 218 | (1.0) | | 42 | (0.8) |  |
| Refused/not stated | 714 | (3.4) | | 255 | (5.0) |  |
| Missing | 3366 | (16.2) | | 455 | (8.9) |  |
| **IMD Quintile (Socioeconomic Status)**** | | | | | | |
| 1- Most deprived | 3886 | (18.8) | | 820 | (16.2) |  |
| 2 | 3959 | (19.2) | | 934 | (18.5) |  |
| 3 | 4208 | (20.4) | | 1017 | (20.1) | <0.0001 |
| 4 | 4312 | (20.9) | | 1111 | (22.0) |  |
| 5- Least Deprived | 4289 | (20.8) | | 1170 | (23.2) |  |

*Chi^2^ test **Patients without postcode, or from Channel Islands or Isle of Man excluded

**Supplementary Table 2: Missing data analysis for individuals missing ethnicity**

|  | **Not missing** | | **Missing** | | P-value* |
| --- | --- | --- | --- | --- | --- |
|  | **n** | **(%)** | **n** | **(%)** |  |
| **IMD Quintile (Socioeconomic Status)** | | | | | |
| 1- Most deprived | 4081 | (18.6) | 625 | (18.6) | 0.001 |
| 2 | 4102 | (18.7) | 791 | (18.7) |  |
| 3 | 4439 | (20.3) | 786 | (20.3) |  |
| 4 | 4608 | (21.0) | 815 | (21.0) |  |
| 5- Least Deprived | 4690 | (21.4) | 769 | (21.4) |  |
| **Age category** | | | | | |
| Paediatric | 1517 | (6.9) | 417 | (10.9) | <0.0001 |
| Adults | 20542 | (93.1) | 3404 | (89.1) |  |
| **Country**** | | | | | |
| England | 20057 | (91.5) | 3498 | (92.4) | <0.0001 |
| Northern Ireland | 103 | (0.5) | 43 | (1.1) |  |
| Scotland | 976 | (4.5) | 131 | (3.5) |  |
| Wales | 784 | (3.6) | 114 | (3.0) |  |

**Supplementary Table 3: Missing data analysis for individuals missing IMD quintile data**

| **Not missing** | | **Missing** | |
| --- | --- | --- | --- |
| n | (%) | n | (%) |
| 25870 | (99.9) | 10 | (0.1) |

**Supplementary Table 4: ERA-EDTA codes and keyword search terms for each Rare Disease Group**

| **Cohort** | **ERA-EDTA code** | **ERA-EDTA code description** | **Keywords** |
| --- | --- | --- | --- |
| **ADPKD** | 2739 | Autosomal dominant (AD) polycystic kidney disease type II | Autosomal dominant polycystic kidney disease, ADPKD, polycystic kidney disease |
|  | 2725 | Autosomal dominant (AD) polycystic kidney disease type I |  |
|  | 2718 | Autosomal dominant (AD) polycystic kidney disease |  |
| **ADTKD** | 2827 | Uromodulin-associated nephropathy (familial juvenile hyperuricaemic nephropathy) | Hyperuricaemic Nephropathy, Primary Hyperuricaemic nephropathy, Familial Hyperuricaemic nephropathy, Medullary cystic kidney disease, Autosomal Dominant Tubulointerstitial Kidney Disease, ADTKD, Familial juvenile hyperuricaemic nephropathy, Familial gouty nephropathy, Familial urate nephropathy, Familial interstitial nephropathy, Uromodulin associated nephropathy, UMOD, Medullary cystic kidney disease type I, Medullary cystic kidney disease type II |
|  | 2804 | Medullary cystic kidney disease type I |  |
|  | 2815 | Medullary cystic kidney disease type II |  |
|  | 1907 | Familial interstitial nephropathy - no histology |  |
|  | 1911 | Familial interstitial nephropathy - histologically proven |  |
| **ARPKD/NPHP** | 2891 | Nephronophthisis - type 6 | Nephronophthisis, NPHP, Autosomal Recessive Polycystic Kidney Disease, ARPKD, Caroli Syndrome with kidney malformation, Caroli Syndrome with cyst, Congenital Hepatic Fibrosis, Nephronophthisis type 1, Nephronophthisis type 2, Nephronophthisis type 3, Nephronophthisis type 4, Nephronophthisis type 5, Nephronophthisis type 6, Senior-Loken, senior–løken, Senior-Loken syndrome, senior–løken syndrome, Joubert, Joubert syndrome, Meckel-Gruber, Meckel-Gruber syndrome, Cogan, Cogan syndrome, Sensenbrenner, Sensenbrenner syndrome, ATD, Asphyxiating thoracic dystrophy, Asphyxiating thoracic dystrophy of the newborn, Chondroectodermal dysplasia-like syndrome, Infantile thoracic dystrophy, JATD, Jeune asphyxiating thoracic dystrophy, Jeune's syndrome, Thoracic pelvic phalangeal dystrophy, Alström, Alström–Hallgren, Alström syndrome, Alström–Hallgren syndrome, Boichis, Senior-Boichis, Boichis syndrome, Senior-Boichis syndrome, Arima, Arima syndrome, Mainzer-Saldino, Mainzer-Saldino syndrome, Bardet-Biedl, Bardet-Biedl syndrome, BBS, Chondroectodermal dysplasia, Ellis Van Creveld syndrome, Mesodermic dysplasia, Mesoectodermal dysplasia |
|  | 2741 | Autosomal recessive (AR) polycystic kidney disease |  |
|  | 2836 | Nephronophthisis |  |
|  | 2858 | Nephronophthisis - type 2 (infantile type) |  |
|  | 2843 | Nephronophthisis - type 1 (juvenile type) |  |
|  | 2870 | Nephronophthisis - type 4 (juvenile type) |  |
|  | 2862 | Nephronophthisis - type 3 (adolescent type) |  |
|  | 2889 | Nephronophthisis - type 5 |  |
| **Alport Syndrome** | 2756 | Alport syndrome - no histology | Alport syndrome, Alport’s syndrome, Alport carrier, Alport, Alport’s, Thin basement membrane nephropathy, TBMN, Autosomal Alport syndrome, Autosomal Alport’s syndrome, X linked Alport syndrome, X linked Alport’s syndrome, X-linked Alport syndrome, X-linked Alport’s syndrome |
|  | 2787 | Thin basement membrane disease |  |
|  | 2760 | Alport syndrome - histologically proven |  |
| **Atypical HUS** | 2668 | Familial haemolytic uraemic syndrome (HUS) | Atypical haemolytic uraemic syndrome, atypical HUS, aHUS, diarrhoea negative HUS, Congenital HUS, Familial HUS, thrombotic microangiopathy, TMA |
|  | 2652 | Congenital haemolytic uraemic syndrome (HUS) |  |
|  | 2623 | Atypical haemolytic uraemic syndrome (HUS) - diarrhoea negative |  |
| **Cystinosis** | 2964 | Cystinosis | Cystinosis |
| **Cystinuria** | 2955 | Cystinuria | Cystinuria |
| **Hyperoxaluria** | 3207 | Primary hyperoxaluria type I | Hyperoxaluria, Primary hyperoxaluria, Oxalosis, Primary Hyperoxaluria Type 1, Primary Hyperoxaluria Type 2, Primary Hyperoxaluria Type 3 |
|  | 3731 | Primary hyperoxaluria type III |  |
|  | 3211 | Primary hyperoxaluria type II |  |
|  | 3194 | Primary hyperoxaluria |  |
|  | 3194 | Primary hyperoxaluria |  |
| **HNF1b mutations** | 1656 | Glomerulocystic Disease | HNF1b, Hepatocyte nuclear factor-1B, Hepatocyte nuclear factor 1B mutation, Hepatocyte nuclear factor 1B, Renal cysts and diabetes, RCAD, Inherited genetic diabetes type 2, MODY 5 |
|  | 1639 | Multicystic Dysplastic Kidneys |  |
|  | 3627 | Renal Cysts & Diabetes Syndrome |  |
|  | 3139 | Diabetes - Type II MODY - Inherited/Genetic |  |
| **Idiopathic Nephrotic Syndrome** | 1100 | Minimal change nephropathy - histologically proven | Primary focal segmental glomerulosclerosis, Primary FSGS, Nephrotic syndrome, NS, INS, Congenital nephrotic syndrome, Congenital NS, Steroid resistant nephrotic syndrome, SRNS, Steroid sensitive nephrotic syndrome, SSRS, Nail Patella Syndrome, Nail-patella syndrome, Denys Drash Syndrome, Denys-Drash Syndrome, Minimal change disease, MCD |
|  | 1349 | Mesangial proliferative glomerulonephritis |  |
|  | 3604 | Nephrotic syndrome of childhood - steroid resistant - no histology |  |
|  | 1090 | Minimal change nephropathy - no histology |  |
|  | 3604 | Nephrotic syndrome of childhood - steroid resistant - no histology |  |
|  | 1090 | Minimal change nephropathy - no histology |  |
|  | 1280 | Familial focal segmental glomerulosclerosis (FSGS) - autosomal recessive - histologically proven |  |
|  | 3615 | Nephrotic syndrome of childhood - no trial of steroids - no histology |  |
|  | 1074 | Denys-Drash syndrome |  |
|  | 1003 | Adult nephrotic syndrome - no histology |  |
|  | 1100 | Minimal change nephropathy - histologically proven |  |
|  | 1279 | Familial focal segmental glomerulosclerosis (FSGS) - autosomal recessive - no histology |  |
|  | 1090 | Minimal change nephropathy - no histology |  |
|  | 1090 | Minimal change nephropathy - no histology |  |
|  | 1100 | Minimal change nephropathy - histologically proven |  |
|  | 1019 | Nephrotic syndrome of childhood - steroid sensitive - no histology |  |
|  | 1100 | Minimal change nephropathy - histologically proven |  |
|  | 1042 | Congenital nephrotic syndrome (CNS) - Finnish type - histologically proven |  |
|  | 1298 | Familial focal segmental glomerulosclerosis (FSGS) - autosomal dominant - no histology |  |
|  | 1035 | Congenital nephrotic syndrome (CNS) - Finnish type - no histology |  |
|  | 1088 | Congenital nephrotic syndrome (CNS) - congenital infection |  |
|  | 1057 | Congenital nephrotic syndrome (CNS) - diffuse mesangial sclerosis |  |
|  | 1026 | Congenital nephrotic syndrome (CNS) - no histology |  |
|  | 1308 | Familial focal segmental glomerulosclerosis (FSGS) - autosomal dominant - histologically proven |  |
|  | 1026 | Congenital nephrotic syndrome (CNS) - no histology |  |
|  | 1061 | Congenital nephrotic syndrome (CNS) - focal segmental glomerulosclerosis (FSGS) |  |
|  | 3253 | Nail-patella syndrome |  |
|  | 1267 | Primary focal segmental glomerulosclerosis (FSGS) |  |
| **IgA Nephropathy** | 1128 | IgA nephropathy - histologically proven | IgA nephropathy, IgA, IgAN |
|  | 1159 | IgA nephropathy secondary to liver cirrhosis - no histology |  |
|  | 1144 | Familial IgA nephropathy - histologically proven |  |
|  | 1137 | Familial IgA nephropathy - no histology |  |
|  | 1163 | IgA nephropathy secondary to liver cirrhosis - histologically proven |  |
| **Inherited Renal Cancer Syndromes** | 3282 | Von Hippel-Lindau disease | Von Hippel Lindau disease, VHL, PTEN hamartoma tumour syndrome, Cowden syndrome, Birt Hogg Dube syndrome, Birt-Hogg-Dube Syndrome, BHD, Hereditary leiomyomatosis and renal cell cancer syndrome, HLRCC, Succinate dehydrogenase-related tumour predisposition syndrome, BAP1-related tumour predisposition syndrome, Hereditary Type 1 papillary renal cell carcinoma syndrome |
| **MGRS** | 2584 | Myeloma cast nephropathy - histologically proven | AH amyloidosis, AHL amyloidosis, C3 glomerulonephritis with monoclonal gammopathy, C3G with monoclonal gammopathy, C3GN with monoclonal gammopathy, Crystalglobulinaemia, Crystal-storing histiocytosis, Fibrillary Glomerulonephritis, Fibrillary GN, Immunotactoid Glomerulonephritis with Organised Microtubular Monoclonal Immunoglobulin Deposits, Immunotactoid GN with Organised Microtubular Monoclonal Immunoglobulin Deposits, GOMMID, Intracapillary monoclonal IgM without cryoglobulin, Intraglomerular lymphoma, Intraglomerular leukaemia, Intracapillary lymphoma, Intracapillary leukaemia, Light chain cast nephropathy, Light chain proximal tubulopathy, crystalline light chain proximal tubulopathy, non crystalline light chain proximal tubulopathy, Monoclonal Immunoglobulin Deposition Disease, MIDD, Light Chain Deposition Disease, LCDD, Heavy Chair Deposition Disease, HCDD, Light and Heavy Chain Deposition Disease, LHCDD, Proliferative glomerulonephritis with monoclonal immunoglobulin deposits, PGNMID, Thrombotic Microangiopathy with monoclonal gammopathy, Type 1 cryoglobulinaemic Glomerulonephritis, Type I cryoglobulinaemic GN, Unclassified MGRS, MGRS |
|  | 2606 | Immunotactoid / fibrillary nephropathy |  |
|  | 2597 | Light chain deposition disease |  |
|  | 2521 | AL amyloid secondary to plasma cell dyscrasia |  |
| **MPGN/C3GN** | 1233 | Mesangiocapillary glomerulonephritis type 2 (dense deposit disease) | Membranoproliferative glomerulonephritis, MPGN, Mesangiocapillary glomerulonephritis, MCGN, Dense Deposit Disease, DDD, C3 Glomerulonephritis, C3 Glomerulopathy, C3GN, C3G, Membranoproliferative glomerulonephritis Type I, MPGN Type I, MPGN I, Membranoproliferative glomerulonephritis Type II, MPGN Type II, MPGN II |
|  | 1222 | Mesangiocapillary glomerulonephritis type 1 |  |
|  | 1246 | Mesangiocapillary glomerulonephritis type 3 |  |
|  | 1233 | Mesangiocapillary glomerulonephritis type 2 (dense deposit disease) |  |
| **Membranous Nephropathy** | 1214 | Membranous nephropathy - infection associated | Membranous nephropathy, Membranous |
|  | 1205 | Membranous nephropathy - drug induced |  |
|  | 1185 | Membranous nephropathy - idiopathic |  |
|  | 1192 | Membranous nephropathy - malignancy associated |  |
| **Pregnancy** | NA | NA | Pregnancy |
| **Retroperitoneal Fibrosis** | 3689 | Retroperitoneal fibrosis secondary to drugs | Retroperitoneal fibrosis, RPF, IgG4-related Vasculitis, IgG4 related vasculitis, IgG4-related disease, IgG4RD, Periaortitis, Aortitis |
|  | 3670 | Retroperitoneal fibrosis secondary to peri-aortitis |  |
|  | 1813 | Idiopathic retroperitoneal fibrosis |  |
| **STEC HUS** | 2610 | Haemolytic uraemic syndrome (HUS) - diarrhoea associated | STEC HUS, STEC-HUS, Shiga toxin associated HUS, Shiga toxin associated Haemolytic Uraemic Syndrome, Shiga toxin veryocytotoxin associated HUS, Shiga toxin veryocytotoxin associated Haemolytic uraemic syndrome |
| **Tuberous Sclerosis Complex** | 3276 | Tuberous sclerosis | Tuberous sclerosis, TS, Tuberous Sclerosis Complex, TSC |
| **Tubulopathy** | 3187 | Familial hypomagnesaemia | Tubulopathy, Dominant hypophosphatemia with nephrolithiasis or osteoporosis, Drug induced Fanconi syndrome, Drug induced hypomagnesemia, Drug induced Nephrogenic Diabetes Insipidus, EAST syndrome, EAST, Epilepsy Ataxia Sensorineural deafness Tubulopathy syndrome, Epilepsy Ataxia Sensorineural deafness Tubulopathy, Familial Hypomagnesaemia with hypercalciuria and nephrocalcinosis, CLDN16/19, Familial primary hypomagnesemia with hypocalcuria, FXYD2, Familial primary hypomagnesemia with normocalcuria, EGF, Familial renal glucosuria, SLC5A2, Fanconi syndrome, Fanconi Renotubular syndrome 1, FRTS1, Fanconi Renotubular syndrome 2, FRTS2, Fanconi Renotubular syndrome 3, FRTS3, Generalized pseudohypoaldosteronism type 1, Heavy metal induced Fanconi syndrome, Heavy metal Fanconi syndrome, Hereditary renal hypouricemia, Hereditary hypophosphatemic rickets with hypercalciuria, Isolated autosomal dominant hypomagnesemia, Glaudemans type, Glaudemans, Glaudeman’s, Liddle syndrome, Liddle, Liddle’s, Nephrogenic diabetes insipidus, Nephrogenic DI, Nephrogenic syndrome of inappropriate antidiuresis, Nephrogenic SIADH, Oncogenic osteomalacia, Osteopetrosis with renal tubular acidosis, Osteopetrosis with RTA, Primary hypomagnesemia with secondary hypocalcemia, Pseudohypoaldosteronism type 2A, Pseudohypoaldosteronism type 2B, Pseudohypoaldosteronism type 2C, Pseudohypoaldosteronism type 2D, Pseudohypoaldosteronism type 2E, Renal pseudohypoaldosteronism type 1, Autoimmune distal renal tubular acidosis, Autoimmune distal RTA, Autosomal dominant distal renal tubular acidosis, Autosomal dominant distal RTA, Autosomal recessive distal renal tubular acidosis, Autosomal recessive distal RTA, Autosomal recessive proximal renal tubular acidosis, Autosomal recessive proximal RTA, Bartter, Bartter syndrome, Bartter Syndrome type 1, Bartter Syndrome type 3 Gitelman Syndrome, Bartter Syndrome Type 4, Familial hypocalciuric hypercalcaemia, Familial hypercalciuric hypocalcaemia, Proximal renal tubular acidosis type II, Proximal RTA type II, Distal renal tubular acidosis type I, Distal RTA type I, Dent and Lowe, Dent & Lowe, Dent disease, Dent’s disease, Lowe syndrome, Oculocerebrorenal syndrome of Lowe, Oculocerebrorenal syndrome, OCRL |
|  | 3187 | Familial hypomagnesaemia |  |
|  | 2901 | Primary Fanconi syndrome |  |
|  | 3156 | Pseudohypoaldosteronism type 2 (Gordon syndrome) |  |
|  | 3156 | Pseudohypoaldosteronism type 2 (Gordon syndrome) |  |
|  | 2901 | Primary Fanconi syndrome |  |
|  | 3085 | Bartter syndrome |  |
|  | 2993 | Hypophosphataemic rickets autosomal recessive (AR) |  |
|  | 2901 | Primary Fanconi syndrome |  |
|  | 3102 | Liddle syndrome |  |
|  | 3044 | Nephrogenic diabetes insipidus |  |
|  | 3156 | Pseudohypoaldosteronism type 2 (Gordon syndrome) |  |
|  | 3037 | Distal renal tubular acidosis with sensorineural deafness - gene mutations |  |
|  | 3141 | Pseudohypoaldosteronism type 1 |  |
|  | 2986 | Hypophosphataemic rickets X-linked (XL) |  |
|  | 3085 | Bartter syndrome |  |
|  | 3156 | Pseudohypoaldosteronism type 2 (Gordon syndrome) |  |
|  | 3092 | Gitelman syndrome |  |
|  | 3085 | Bartter syndrome |  |
|  | 3187 | Familial hypomagnesaemia |  |
|  | 3160 | Familial hypocalciuric hypercalcaemia |  |
|  | 2972 | Inherited renal glycosuria |  |
|  | 3173 | Familial hypercalciuric hypocalcaemia |  |
|  | 3028 | Distal renal tubular acidosis (RTA) - type I |  |
|  | 3016 | Proximal renal tubular acidosis (RTA) - type II |  |
|  | 2917 | Tubular disorder as part of inherited metabolic diseases |  |
|  | 3156 | Pseudohypoaldosteronism type 2 (Gordon syndrome) |  |
|  | 2929 | Dent disease |  |
|  | 2938 | Lowe syndrome (oculocerebrorenal syndrome) |  |
| **ANCA positive vasculitis** | 1383 | Systemic vasculitis - ANCA negative - histologically proven | MPO Vasculitis, MPO-Vasculitis, PR3 vasculitis, PR3-vasculitis, Granulomatosis with polyangiitis, Polyangiitis, GPA, Wegener, Wegener’s, Eosinophilic granulomatosis with polyangiitis, EGPA, Churg Strauss, ANCA Vasculitis |
|  | 1429 | Microscopic polyangiitis - histologically proven |  |
|  | 3852 | Systemic vasculitis - ANCA positive - histologically proven |  |
|  | 1401 | Granulomatosis with polyangiitis - no histology |  |
|  | 1417 | Granulomatosis with polyangiitis - histologically proven |  |
|  | 1396 | Systemic vasculitis - ANCA positive - no histology |  |
|  | 1438 | Churg-Strauss syndrome - no histology |  |
|  | 1440 | Churg-Strauss syndrome - histologically proven |  |
| **Anti-GBM disease** | 1472 | Anti-Glomerular basement membrane (GBM) disease / Goodpasture's syndrome - histologically proven | Anti GBM disease, Anti-GBM disease, Anti GBM, Anti-GBM, Anti Glomerular basement membrane, Anti-Glomerular Basement, |
|  | 1464 | Anti-Glomerular basement membrane (GBM) disease / Goodpasture's syndrome - no histology |  |
| **Other Vasculitides** | 3847 | Systemic vasculitis - ANCA negative - no histology | Vasculitis, Small vessel vasculitis, IgA vasculitis, IgAV, Henoch Schonlein purpura, Henoch Schonlein, Henoch Schönlein, HSP, Cryoglobulinaemic vasculitis, Classical Polyarteritis Nodosa, Polyarteritis Nodosa, PAN, Kawasaki disease, Kawasaki, Kawasaki’s, Giant cell arteritis, GCA, Takayasu’s arteritis, Takayasu, Takayasu’s, Behcet’s disease, Behcet’s, Behcet, Cogan’s syndrome, Cogan’s, Cogan, Isolated aortitis, Primary cerebral aortitis |
|  | 1515 | Henoch-Schönlein purpura / nephritis - histologically proven |  |
|  | 1504 | Henoch-Schönlein purpura / nephritis - no histology |  |
|  | 1455 | Polyarteritis nodosa |  |

**Supplementary Table 5: Ethnicity and IMD Quintile*, stratified by current age**

| **Ethnicity** | **White** | | **Mixed** | | **Asian** | | **Black** | | **Other** | | **P-value**** |
| --- | --- | --- | --- | --- | --- | --- | --- | --- | --- | --- | --- |
|  | n | (%) | n | (%) | n | (%) | n | (%) | n | (%) |  |
| **Paediatric** | 1103 | (74.7) | 49 | (3.3) | 255 | (17.3) | 48 | (3.3) | 21 | (1.4) | <0.0001 |
| **Adults** | 17185 | (87.6) | 149 | (0.8) | 1471 | (7.5) | 570 | (2.9) | 239 | (1.2) |  |
| **IMD Quintile** | **1- Most deprived** | | **2** | | **3** | | **4** | | **5- Least deprived** | | **P-value**** |
|  | n | (%) | n | (%) | n | (%) | n | (%) | n | (%) |  |
| **Paediatric** | 580 | (30.3) | 318 | (16.6) | 342 | (17.9) | 341 | (17.8) | 334 | (17.4) | <0.0001 |
| **Adults** | 4126 | (17.3) | 4575 | (19.2) | 4883 | (20.5) | 5082 | (21.4) | 5125 | (21.5) |  |

*Complete case analysis **Chi^2^ test

**Supplementary Table 6: Ethnic and IMD Quintile distribution of children (aged ≤18 years) in England* compared to the children in RaDaR**

|  | **Children in the English population** | | **Children in RaDaR population** | | | **P-value**** |
| --- | --- | --- | --- | --- | --- | --- |
|  | n | (%) | N | | (%) |  |
| **Ethnicity** | | | | | | |
| White | 9495175 | (79) | 1008 | (73) | | <0.0001 |
| Mixed | 603396 | (5) | 48 | (4) | |  |
| Asian | 1155283 | (10) | 249 | (18) | |  |
| Black | 567083 | (5) | 47 | (3) | |  |
| Other | 149430 | (1) | 20 | (1) | |  |
| Total | 11970367 | (100) | 1372 | (100) | |  |
| **IMD Quintile** | | | | | | |
| 1-Most deprived | 3151456 | (24) | 548 | (31) | | <0.0001 |
| 2 | 2753699 | (21) | 298 | (17) | |  |
| 3 | 2514215 | (19) | 320 | (18) | |  |
| 4 | 2402419 | (18) | 318 | (18) | |  |
| 5- Least deprived | 2460532 | (19) | 305 | (17) | |  |
| Total | 13282321 | (100) | 1,789 | (100) | |  |

*Office of National Statistics 2011 census data **Chi2 test

**Supplementary Table 7: Monogenic and Non monogenic disorders, stratified by ethnicity**

|  | **Monogenic** | | **Non monogenic** | | **P-value** |
| --- | --- | --- | --- | --- | --- |
|  | n | (%) | N | (%) |  |
| White | 6886 | (90.3) | 11486 | (84.7) | <0.0001 |
| Mixed | 70 | (0.9) | 128 | (0.9) |  |
| Asian | 400 | (5.2) | 1333 | (9.8) |  |
| Black | 187 | (2.5) | 435 | (3.2) |  |
| Other | 83 | (1.1) | 177 | (1.3) |  |

**Chi^2^ test . Excluding patients with ethnicity data missing. Patients with two diagnoses are included for each diagnosis.

|  | | **Monogenic** | | **Non monogenic** | | **P-value**** |
| --- | --- | --- | --- | --- | --- | --- |
|  |  | n | (%) | n | (%) |  |
| **All** | Paediatric | 575 | (29.7) | 1360 | (70.3) | <0.0001 |
|  | Adults | 9352 | (38.9) | 14717 | (61.1) |  |
| **White** | Paediatric | 311 | (28.2) | 793 | (71.8) | <0.0001 |
|  | Adults | 6575 | (38.1) | 10693 | (61.9) |  |
| **Mixed** | Paediatric | 11 | (22.4) | 38 | (77.6) | 0.029 |
|  | Adults | 59 | (39.6) | 90 | (60.4) |  |
| **Asian** | Paediatric | 61 | (23.9) | 194 | (76.1) | 0.730 |
|  | Adults | 339 | (22.9) | 1139 | (77.1) |  |
| **Black** | Paediatric | 8 | (16.7) | 40 | (83.3) | 0.035 |
|  | Adults | 179 | (31.2) | 395 | (68.8) |  |
| **Other** | Paediatric | 7 | (33.3) | 14 | (66.7) | 0.885 |
|  | Adults | 76 | (31.8) | 163 | (68.2) |  |

**Supplementary Table 8: Monogenic and Non monogenic disorders, stratified by age category* and ethnicity**

*Age on July 25^th^ 2022 **Chi^2^ test. Excluding patients with ethnicity data missing. Patients with two diagnoses are included for each diagnosis.

**Supplementary Table 9: Monogenic and Non monogenic disorders, stratified by Ethnicity and IMD Quintile**

|  | | **1- Most deprived** | | **2** | | **3** | | **4** | | **5- Least deprived** | | **P-value*** |
| --- | --- | --- | --- | --- | --- | --- | --- | --- | --- | --- | --- | --- |
|  |  | n | (%) | n | (%) | n | (%) | n | (%) | n | (%) |  |
| **All** | Monogenic | 1671 | (16.9) | 1878 | (19.0) | 2063 | (20.9) | 2122 | (21.5) | 2129 | (21.6) | <0.0001 |
|  | Not Monogenic | 3066 | (19.2) | 3039 | (19.0) | 3185 | (19.9) | 3320 | (20.8) | 3357 | (21.0) |  |
| **White** | Monogenic | 1099 | (16.0) | 1213 | (17.7) | 1445 | (21.1) | 1524 | (22.3) | 1567 | (22.9) | 0.429 |
|  | Not Monogenic | 1948 | (17.1) | 2003 | (17.6) | 2331 | (20.5) | 2534 | (22.2) | 2578 | (22.6) |  |
| **Mixed** | Monogenic | 14 | (20.0) | 20 | (28.6) | 12 | (17.1) | 12 | (17.1) | 12 | (17.1) | 0.718 |
|  | Not Monogenic | 32 | (25.0) | 31 | (24.2) | 24 | (18.8) | 15 | (11.7) | 26 | (20.3) |  |
| **Asian** | Monogenic | 145 | (36.5) | 83 | (20.9) | 64 | (16.1) | 56 | (14.1) | 49 | (12.3) | 0.826 |
|  | Not Monogenic | 451 | (33.9) | 312 | (23.5) | 214 | (16.1) | 185 | (13.9) | 168 | (12.6) |  |
| **Black** | Monogenic | 77 | (41.4) | 48 | (25.8) | 32 | (17.2) | 16 | (8.6) | 13 | (7.0) | 0.254 |
|  | Not Monogenic | 158 | (36.3) | 150 | (34.5) | 74 | (17.0) | 32 | (7.4) | 21 | (4.8) |  |
| **Other** | Monogenic | 12 | (14.5) | 25 | (30.1) | 21 | (25.3) | 13 | (15.7) | 12 | (14.5) | 0.010 |
|  | Not Monogenic | 57 | (32.2) | 42 | (23.7) | 27 | (15.3) | 17 | (9.6) | 34 | (19.2) |  |

*Chi^2^ test. Excluding patients with ethnicity and IMD Quintile data missing. Patients with two diagnoses are included for each diagnosis.

**Supplementary Table 10: Socioeconomic status (IMD Quintile) of RaDaR participants, stratified by mode of inheritance and age category***

|  | | **1- Most deprived** | | **2** | | **3** | | **4** | | **5- Least deprived** | | **P-value**** |
| --- | --- | --- | --- | --- | --- | --- | --- | --- | --- | --- | --- | --- |
|  |  | **n** | **(%)** | **n** | **(%)** | **n** | **(%)** | **n** | **(%)** | **n** | **(%)** |  |
| **Autosomal dominant** | | 1263 | (16.1) | 1485 | (18.9) | 1643 | (20.9) | 1721 | (21.9) | 1747 | (22.2) | <0.0001 |
| **Autosomal Recessive/X-linked** | | 408 | (20.4) | 393 | (19.6) | 420 | (21.0) | 401 | (20.0) | 382 | (19.1) |  |
| **Mostly non monogenic** | | 3066 | (19.2) | 3039 | (19.0) | 3185 | (19.9) | 3320 | (20.8) | 3357 | (21.0) |  |
| **Autosomal Dominant** | Paediatric | 62 | (28.2) | 38 | (17.3) | 33 | (15.0) | 42 | (19.1) | 45 | (20.5) | <0.0001 |
|  | Adults | 1201 | (15.7) | 1447 | (18.9) | 1610 | (21.1) | 1679 | (22.0) | 1702 | (22.3) |  |
| **Autosomal Recessive/X-linked** | Paediatric | 112 | (32.0) | 59 | (16.9) | 65 | (18.6) | 63 | (18.0) | 51 | (14.6) | <0.0001 |
|  | Adults | 296 | (17.9) | 334 | (20.2) | 355 | (21.5) | 338 | (20.4) | 331 | (20.0) |  |
| **Mostly non monogenic** | Paediatric | 406 | (30.2) | 222 | (16.5) | 244 | (18.1) | 236 | (17.5) | 238 | (17.7) | <0.0001 |
|  | Adults | 2660 | (18.2) | 2817 | (19.3) | 2941 | (20.1) | 3084 | (21.1) | 3119 | (21.3) |  |

*age on 25^th^ July 2022 **Chi^2^ test

**Supplementary Table 11: IMD Quintile, stratified by current age and Ethnicity**

|  |  | 1- Most deprived | | 2 | | 3 | | 4 | | 5- Least deprived | | p-value* |
| --- | --- | --- | --- | --- | --- | --- | --- | --- | --- | --- | --- | --- |
|  |  | n | (%) | n | (%) | n | (%) | n | (%) | n | (%) |  |
| White | Paediatric | 272 | (24.8) | 183 | (16.7) | 207 | (18.9) | 213 | (19.4) | 221 | (20.2) | p<0.0001 |
|  | Adults | 2758 | (16.2) | 3017 | (17.7) | 3550 | (20.8) | 3831 | (22.5) | 3906 | (22.9) |  |
| Mixed | Paediatric | 10 | (33.3) | 10 | (33.3) | ≤6 | NR** | ≤6 | NR | 10 | (33.3) | p=0.626 |
|  | Adults | 33 | (22.1) | 39 | (26.2) | 30 | (20.1) | 21 | (14.1) | 26 | (17.4) |  |
| Asian | Paediatric | 136 | (53.5) | 46 | (18.1) | 26 | (10.2) | 25 | (9.8) | 21 | (8.3) | p<0.0001 |
|  | Adults | 456 | (31.1) | 347 | (23.7) | 251 | (17.1) | 216 | (14.7) | 196 | (13.4) |  |
| Black | Paediatric | 25 | (55.5) | 10 | (22.2) | 10 | (22.2) | ≤6 | NR | ≤6 | NR | p=0.489 |
|  | Adults | 208 | (36.6) | 186 | (32.7) | 98 | (17.2) | 45 | (7.9) | 32 | (5.6) |  |
| Other | Paediatric | 10 | (50.0) | 10 | (50.0) | ≤6 | NR | ≤6 | NR | 0 | (0.0) | 0.017 |
|  | Adults | 59 | (24.7) | 59 | (24.7) | 46 | (19.2) | 29 | (12.1) | 46 | (19.2) |  |

*Fishers exact test **NR- Not reported; cells with fewer than 6 patients not reported due to risk of re-identification. Where cells are not reported, corresponding cell values are rounded to nearest 5

**Supplementary Table 12: Socioeconomic status (IMD Quintile), stratified by monogenic and non monogenic disorders and age category***

|  | | **1- Most deprived** | | **2** | | **3** | | **4** | | **5- Least deprived** | | **P-value**** |
| --- | --- | --- | --- | --- | --- | --- | --- | --- | --- | --- | --- | --- |
|  |  | n | (%) | n | (%) | n | (%) | n | (%) | n | (%) |  |
| **Paediatric** | Monogenic | 174 | (30.5) | 97 | (17.0) | 98 | (17.2) | 105 | (18.4) | 96 | (16.8) | 0.961 |
|  | Non monogenic | 406 | (30.2) | 222 | (16.5) | 244 | (18.1) | 236 | (17.5) | 238 | (17.7) |  |
| **Adults** | Monogenic | 1497 | (16.1) | 1781 | (19.2) | 1965 | (21.1) | 2017 | (21.7) | 2033 | (21.9) | 0.001 |
|  | Non monogenic | 2660 | (18.2) | 2817 | (19.3) | 2941 | (20.1) | 3084 | (21.1) | 3119 | (21.3) |  |
| **Monogenic** | Paediatric | 174 | (30.5) | 97 | (17.0) | 98 | (17.2) | 105 | (18.4) | 96 | (16.8) | <0.0001 |
|  | Adults | 1497 | (16.1) | 1781 | (19.2) | 1965 | (21.1) | 2017 | (21.7) | 2033 | (21.9) |  |
| **Non monogenic** | Paediatric | 406 | (30.2) | 222 | (16.5) | 244 | (18.1) | 236 | (17.5) | 238 | (17.7) | <0.0001 |
|  | Adults | 2660 | (18.2) | 2817 | (19.3) | 2941 | (20.1) | 3084 | (21.1) | 3119 | (21.3) |  |

*Age on July 25^th^ 2022 **Chi^2^ test. Excluding patients with ethnicity data missing. Patients with two diagnoses are included for each diagnosis.

Supplementary Table 13: Number and percentage of rare disease diagnoses for each disorder within UKRR and RaDaR KRT recipients

| **Rare Disease Group** | **RaDaR KRT population** | | **UKRR rare disease KRT population** | |
| --- | --- | --- | --- | --- |
|  | **n** | **(%)** | **n** | **(%)** |
| ADPKD | 2986 | (35.1) | 6880 | (32.4) |
| ADTKD | 88 | (1.0) | 318 | (1.5) |
| ARPKD/NPHP | 99 | (1.2) | 237 | (1.1) |
| Alport Syndrome | 344 | (4.0) | 716 | (3.4) |
| Cystinosis | 87 | (1.0) | 133 | (0.6) |
| Cystinuria | 7 | (0.1) | 11 | (0.1) |
| Hyperoxaluria | 36 | (0.4) | 76 | (0.4) |
| INS | 666 | (7.8) | 2324 | (11) |
| IgA Nephropathy | 2109 | (24.8) | 5772 | (27.2) |
| MGRS | 76 | (0.9) | 275 | (1.3) |
| MPGN/C3GN | 476 | (5.6) | 825 | (3.9) |
| Membranous Nephropathy | 425 | (5.0) | 929 | (4.4) |
| Renal cancer inherited | ≤6 | (0.0) | 15 | (0.1) |
| Retroperitoneal Fibrosis | 15 | (0.2) | 29 | (0.1) |
| STEC HUS | 28 | (0.3) | 60 | (0.3) |
| Tuberous Sclerosis | 21 | (0.2) | 33 | (0.2) |
| Tubulopathies | 28 | (0.3) | 72 | (0.3) |
| Vasculitis | 904 | (10.6) | 2201 | (10.4) |
| aHUS | 122 | (1.4) | 297 | (1.4) |
| Total Rare Disease KRT Recipients | 8517 | (100) | 21203 | (100) |

* KRT = Kidney replacement therapy, UKRR = UK Renal Registry, denominator being the total number of KRT recipients with an eligible rare disease diagnosis

**Supplementary Figure 1: Comparison of ethnicity in each Rare Disease Group to total ethnicity breakdown of RaDaR**

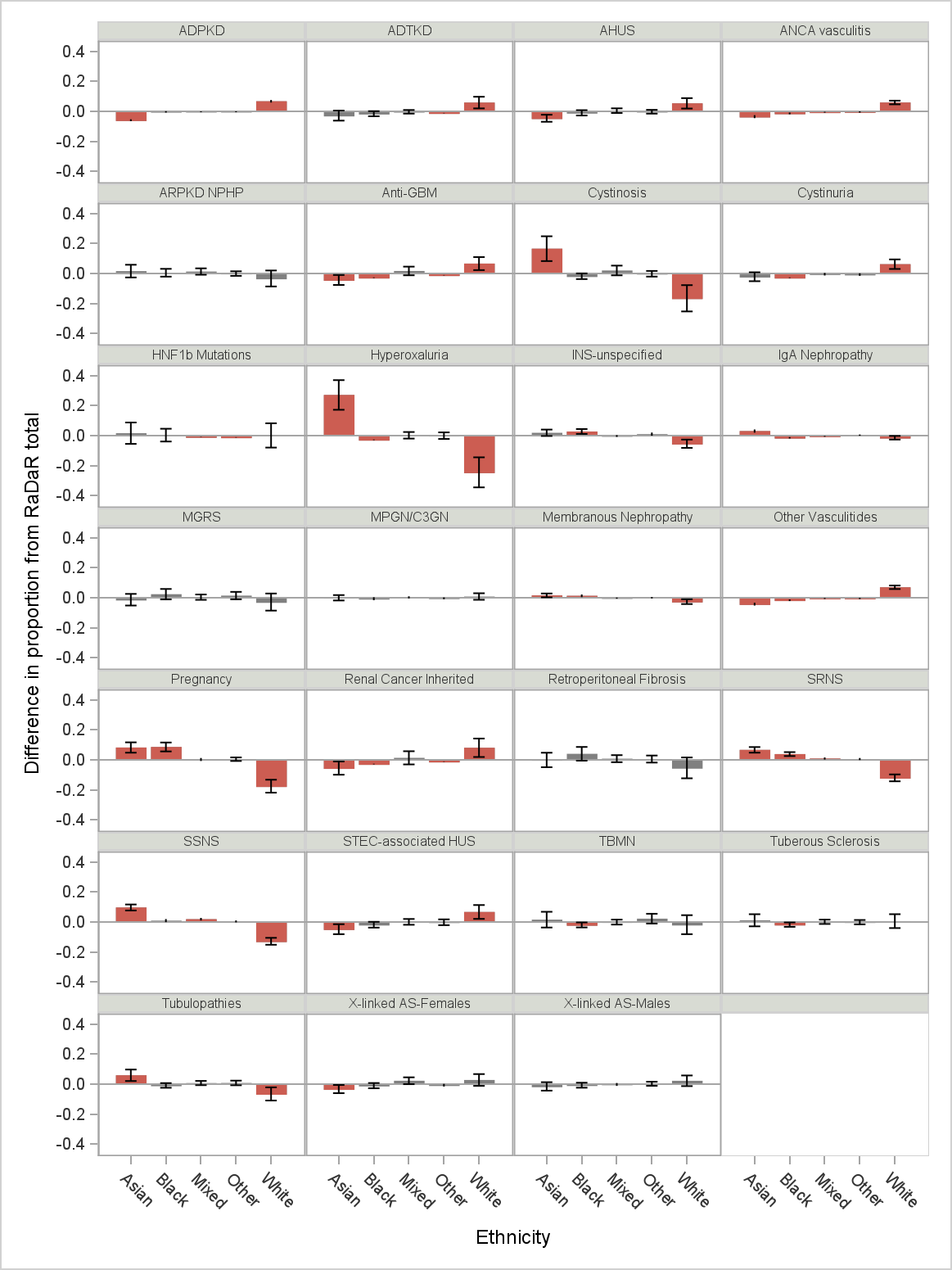

Proportion of patients of each ethnicity for each RDG compared to the ethnicity breakdown of RaDaR using the two-sample test of proportions (Z-test). Bars highlighted in red where Z-test p value <0.05.

**Supplementary Figure 2: Comparison of ethnicity in each Rare Disease Group to total ethnicity breakdown of RaDaR, stratified by current a) paediatric b) adult patients**

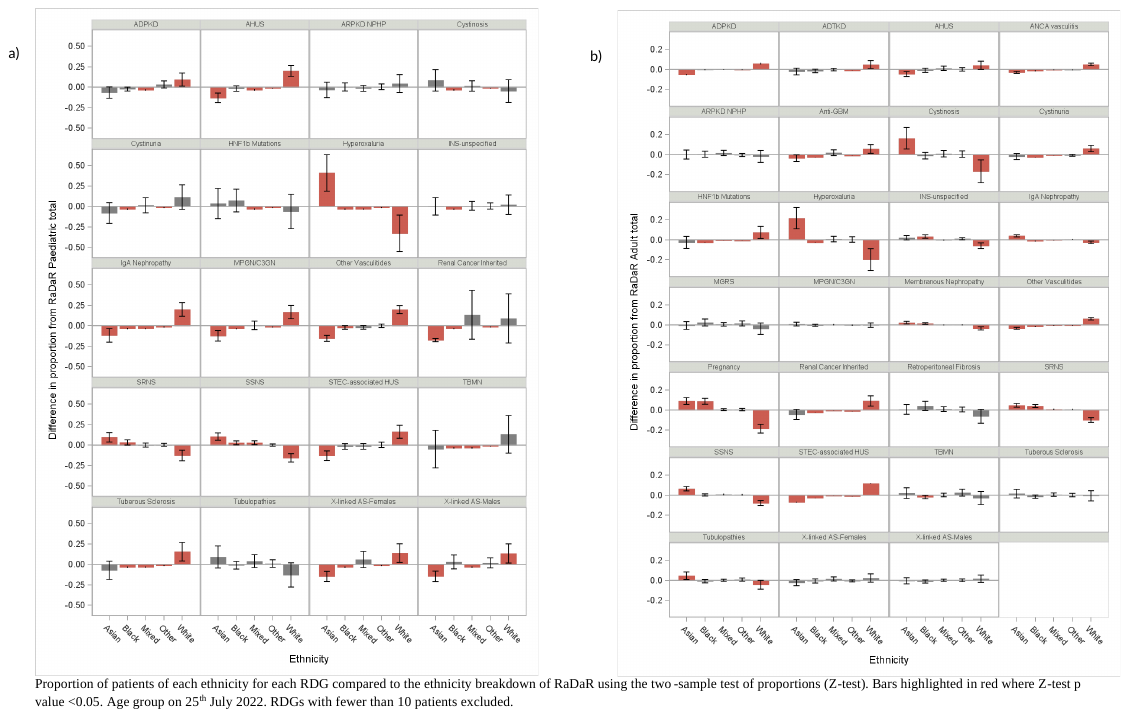

**Supplementary Figure 3: Comparison of IMD Quintile in each Rare Disease Group to total IMD Quintile distribution of RaDaR**

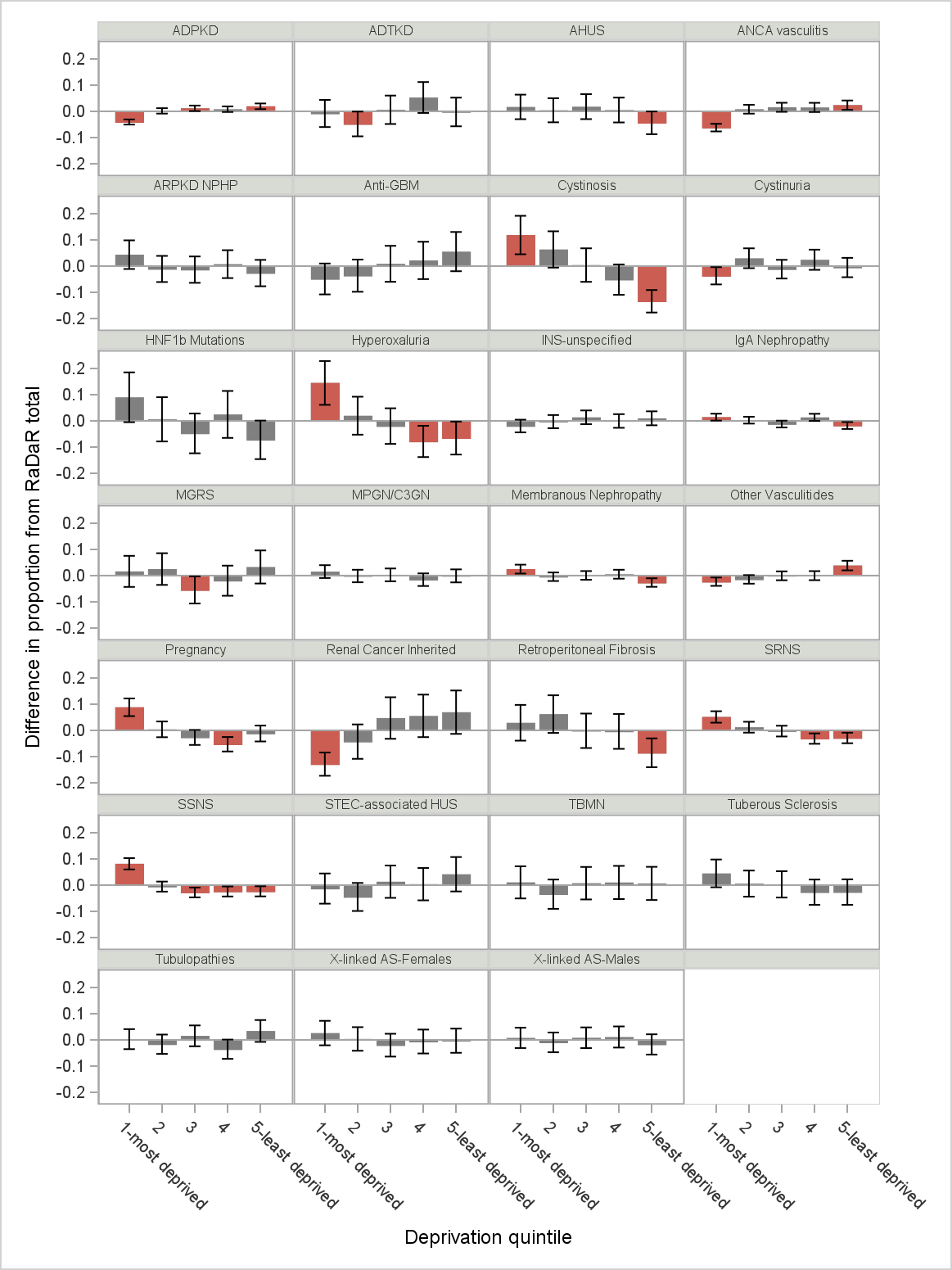
Proportion of patients in each IMD Quintile for each RDG compared to the IMD Quintile breakdown of RaDaR using the two-sample test of proportions (Z-test). Bars highlighted in red where Z-test p value <0.05.

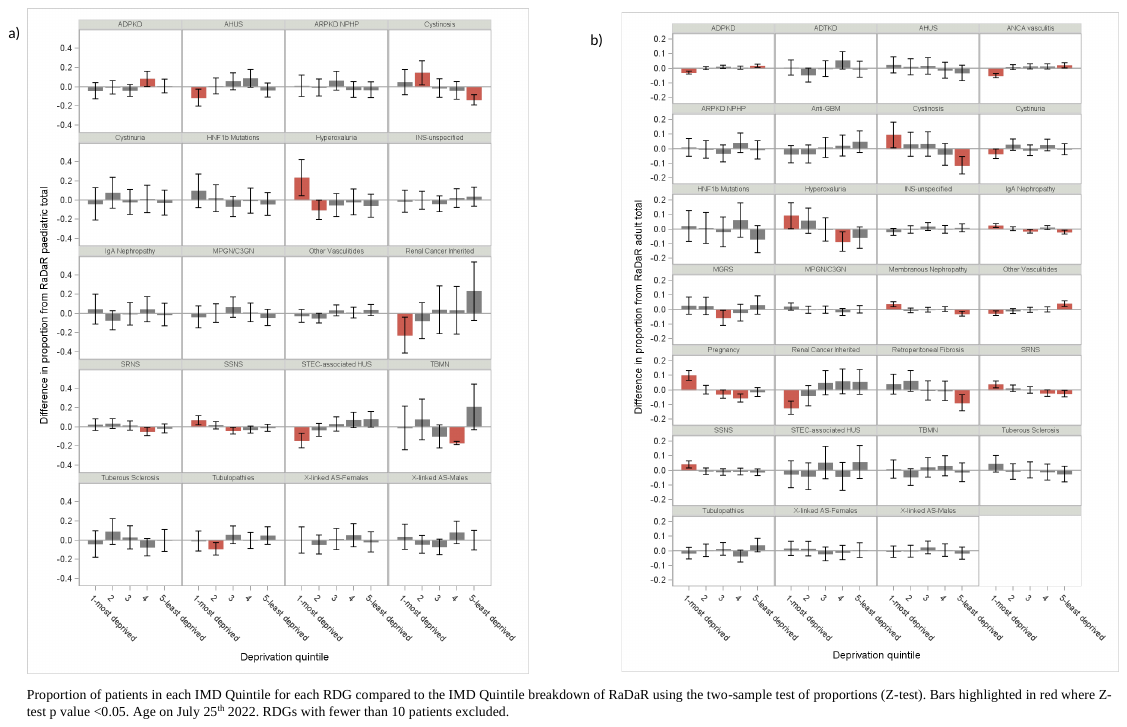
**Supplementary Figure 4: Comparison of IMD Quintile in each Rare Disease Group to total IMD Quintile distribution of RaDaR, stratified by current a) paediatric b) adult patients**

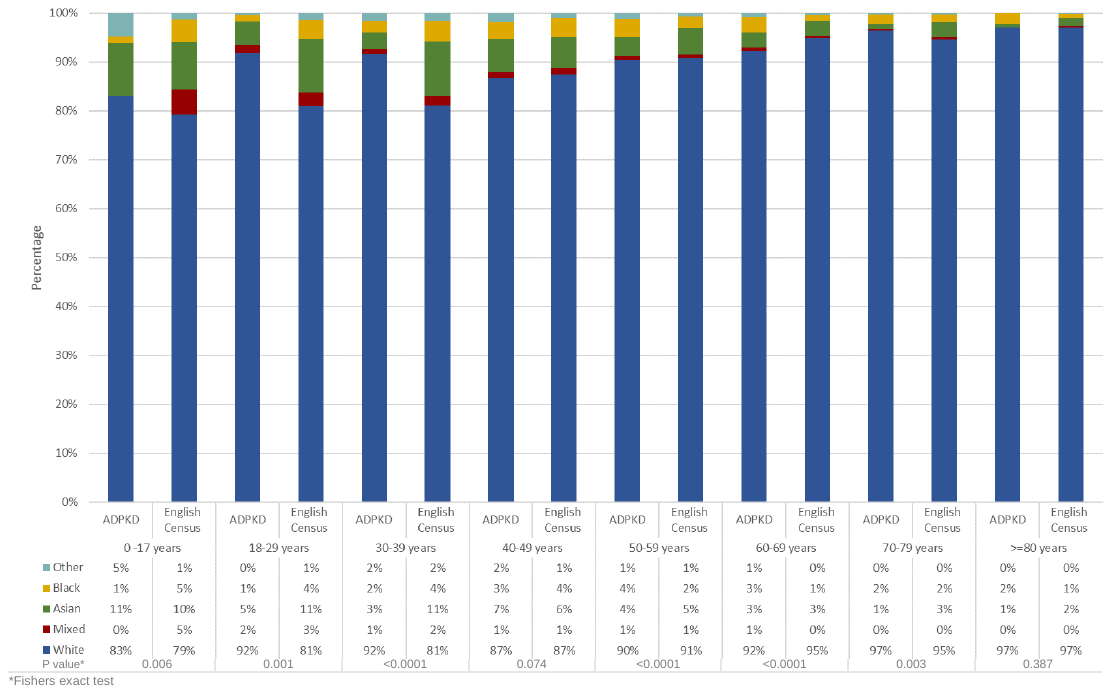
**Supplementary Figure 5: Ethnicity of English RaDaR patients with ADPKD compared to English census**
