## Supplementary appendix 1 Radar consortium for "The National Registry of Rare Kidney Diseases (RaDaR): Description, recruitment, and cross-sectional analyses of 25,880 adults and children with rare kidney diseases in the UK"

Sharirose Abat[1], Shazia Adalat[2], Joy Agbonmwandolor[3], Zubaidah Ahmad[4], Abdulfattah Alejmi[5], Rashid Almasarwah[6], Nicholas Annear[1], Ellie Asgari[4], Amanda Ayers[7], Jyoti Baharani[8], Gowrie Balasubramaniam[9], Felix Jo-Bamba Kpodo[10], Tarun Bansal[11], Alison Barratt[12], Jonathan Barratt[79], Megan Bates[13], Natalie Bayne[14], Janet Bendle[15], Sarah Benyon[16], Carsten Bergmann[17, 18], Sunil Bhandari[19], Coralie Bingham[20], Preetham Boddana[21], Sally Bond[22], Fiona Braddon[23], Kate Bramham[23], Angela Branson[15], Stephen Brearey[24], Vicky Brocklebank[25], Sharanjit Budwal[26], Conor Byrne[27], Hugh Cairns[28], Brian Camilleri[29], Gary Campbell[30], Alys Capell[31], Margaret Carmody[8], Marion Carson[32], Tracy Cathcart[19], Christine Catley[9], Karine Cesar[33], Houda Chea[15], James Chess[34], Chee Kay Cheung[26], Katy-Jane Chick[35], Nihil Chitalia[36], Martin Christian[37], Tina Chrysochou[38, 39], Katherine Clark[40], Christopher Clayton[41], Rhian Clissold[20], Helen Cockerill[33], Joshua Coelho[42], Elizabeth Colby[43], Viv Colclough[44], Eileen Conway[45], H. Terence Cook[46], Wendy Cook[47], Theresa Cooper[48], Richard J Coward[43], Sarah Crosbie[22], Gabor Cserep[49], Anjali Date[50], Katherine Davidson[48], Amanda Davies[51], Neeraj Dhaun[52], Ajay Dhaygude[53], Lynn Diskin[12], Abhijit Dixit[41, 54], Eunice Ann Doctolero[35], Suzannah Dorey[55], Lewis Downard[23], Mark Drayson[56], Gavin Dreyer[27], Tina Dutt[57], Kufreabasi Etuk[28], Dawn Evans[58], Jenny Finch[29], Frances Flinter[59], James Fotheringham[60], Daniel P. Gale[61], Hugh Gallagher[62], David Game[4], Eva Lozano Garcia[42], Madita Gavrila[22], Susie Gear[63], Colin Geddes[64], Mark Gilchrist[65], Matt Gittus[66], Paraskevi Goggolidou[67], Christopher Goldsmith[57], Patricia Gooden[68], Andrea Goodlife[26], Priyanka Goodwin[53], Tassos Grammatikopoulos[28, 69], Barry Gray[70], Megan Griffith[46], Steph Gumus[9], Sanjana Gupta[71], Patrick Hamilton[72], Lorraine Harper[56], Tess Harris[73], Louise Haskell[74], Samantha Hayward[43], Shivaram Hegde[75], Bruce Hendry[76], Sue Hewins[77], Nicola Hewitson[78], Kate Hillman[15], Mrityunjay Hiremath[57], Alexandra Howson[79], Zay Htet[28], Sharon Huish[16], Richard Hull[1], Alister Humphries[68], David P. J. Hunt[119], Karl Hunter[80], Samantha Hunter[19], Marilyn Ijeomah-Orji[6], Nick Inston[81], David Jayne[82], Gbemisola Jenfa[31], Alison Jenkins[83], Sally Johnson[118], Caroline A Jones[84], Colin Jones[85], Amanda Jones[5], Lavanya Kamesh[81], Durga Kanigicherla[39], Fiona Karet Frankl[82], Mahzuz Karim[86], Amrit Kaur[87], David Kavanagh[25], Kelly Kearley[88], Larissa Kerecuk[14], Arif Khwaja[70], Garry King[23], Grant King[89], Ewa Kislowska[4], Edyta Klata[29], Maria Kokocinska[14], Mark Lambie[90], Laura Lawless[41], Thomas Ledson[80], Rachel Lennon[91], Adam P Levine[92], Ling Wai Maggie Lai[16], Graham Lipkin[81], Graham Lovitt[93], Paul Lyons[94], Holly Mabillard[95], Katherine Mackintosh[7], Khalid Mahdi[96], Eamonn Maher[97], Kevin J. Marchbank[25], Patrick B Mark[64], Bridgett Masunda[9], Zainab Mavani[31], Jake Mayfair[4], Joanna Mckinnell[98], Nabil Melhem[2], Simon Meyrick[51], Shabbir Moochhala[61], Putnam Morgan[99], Ann Morgan[100, 101], Fawad Muhammad[5], Shona Murray[30], Kristina Novobritskaya[22], Albert CM Ong[66, 70], Louise Oni[102], Kate Osmaston[23], Neal Padmanabhan[64], Sharon Parkes[14], Jean Patrick[7], James Pattison[4], Riny Paul[1], Rachel Percival[103], Stephen J. Perkins[104], Alexandre Persu[105, 106], William G Petchey[107], Matthew C. Pickering[46], Jennifer Pinney[81], David Pitcher[23], Lucy Plumb[43], Zoe Plummer[23], Joyce Popoola[1], Frank Post[28], Albert Power[83], Guy Pratt[56], Charles Pusey[46], Ria Rabara[22], May Rabuya[4],Tina Raju[42], Chadd Javier[108], Ian SD Roberts[22], Candice Roufosse[109], Adam Rumjon[28], Moin Saleem[43], RN Sandford[97], Kanwaljit S. Sandu[110], Nadia Sarween[81], John A. Sayer[95], Neil Sebire[111, 112], Haresh Selvaskandan[26], Asheesh Sharma[57], Edward J Sharples[22], Neil Sheerin[25], Harish Shetty[53], Rukshana Shroff[112], Roslyn Simms[70], Manish Sinha[2], Smeeta Sinha[113], Kerry Smith[29], Lara Smith[15], Shalabh Srivastava[114], Retha Steenkamp[23], Ian Stott[115], Katerina Stroud[97], Pauline Swift[42], Justyna Szklarzewicz[26], Fred Tam[46], Kay Tan[116], Robert Taylor[117], Marc Tischkowitz[97], Kay Thomas[4], Yincent Tse[118], Alison Turnbull[85], A Neil Turner[119], Kay Tyerman[55], Miranda Usher[120], Gopalakrishnan Venkat-Raman[121], Alycon Walker[122], Stephen B. Walsh[61], Aoife Waters[123], Angela Watt[68], Phil Webster[6], Ashutosh Wechalekar[124], Gavin Iain Welsh[43], Nicol West[125], David Wheeler[61], Lisa Willcocks[107,] Angharad Williams[33], Emma Williams[29], Karen Williams[4], Deborah H Wilson[126], Patricia D Wilson[127],

Paul Winyard[9], Edwin Wong[25], Katie Wong[23], Grahame Wood[58], Emma Woodward[15], Len Woodward[128], Adrian Woolf[129], David Wright[71]

[1]St George's University Hospitals NHS Foundation Trust, UK

[2]Evelina London Children's Hospital, UK

[3]David Evans Medical Research Centre, Nottingham University Hospital NHS Trust, UK

[4]Guy's and St Thomas NHS foundation Trust, UK

[5]Ysbyty Gwynedd, Betsi Cadwaladr University Health Board, UK

[6]Imperial College Healthcare NHS Trust, UK

[7]James Paget University Hospital NHS Foundation Trust, UK

[8]Heart of England NHS Foundation Trust, Birmingham, UK

[9]Mid and South Essex NHS Foundation Trust, UK

[10]Royal Berkshire NHS Foundation Trust, UK

[11]Bradford Teaching Hospitals NHS Foundation Trust, UK

[12]Royal United Hospital Bath NHS Trust, UK

[13]Freeman Hospital, Newcastle Upon Tyne, UK

[14]Birmingham Women's and Children's NHS Foundation Trust, UK

[15]Manchester University NHS Foundation Trust, UK

[16]Royal Devon University Healthcare NHS Foundation Trust, UK

[17]Medizinische Genetik Mainz, Mainz, Germany

[18]Department of Medicine, Faculty of Medicine, Medical Center-University of Freiburg, Freiburg, Germany

[19]Hull University Teaching Hospitals NHS Trust, UK

[20]Exeter Kidney Unit, Royal Devon University Healthcare NHS Foundation Trust, UK

[21]Gloucestershire Hospitals NHS Foundation Trust, UK

[22]Oxford University Hospitals NHS Foundation Trust, UK

[23]UK Kidney Association, UK

[24]Countess of Chester NHS Foundation Trust, UK

[25]National Renal Complement Therapeutics Centre, Newcastle upon Tyne Hospitals NHS Foundation Trust, Newcastle upon Tyne, UK

[26]University Hospitals of Leicester NHS Trust, UK

[27]Barts Health NHS Trust, London, UK

[28]King's College Hospital NHS Foundation Trust, UK

[29]East Suffolk and North Essex NHS Foundation Trust, UK

[30]Ninewells Hospital and Medical School, Dundee, UK

[31]North West Anglia NHS Foundation Trust, UK

[32]Northern Health and Social Care Trust and Northern Ireland Clinical Research Network

[33]West Suffolk NHS Foundation Trust, UK

[34]Morriston Hospital, Swansea Bay Health Board, UK

[35]Lister Hospital, East and North Hertfordshire NHS Trust, UK

[36]Dartford and Gravesham NHS Trust, UK

[37]Nottingham Children's Hospital, UK

[38]Salford Royal Hospital, Northern Care Alliance NHS Foundation Trust, Salford, UK

[39]University of Manchester, UK

[40]King's College London, UK

[41]Nottingham University Hospitals NHS trust, UK

[42]Epsom and St Helier University Hospitals NHS Trust, UK

[43]University of Bristol Medical School, Bristol, UK

[44]Royal Stoke University Hospital, UK

[45]Manchester Royal Infirmary, UK

[46]Centre for Inflammatory Disease, Imperial College London, UK

[47]Nephrotic Syndrome Trust' (NeST) , UK

[48]North Cumbria Integrated Care NHS Foundation Trust, UK

[49]Colchester General Hospital, UK

[50]Tameside and Glossop Integrated Care NHS Foundation Trust, UK

[51]Wye Valley NHS Trust, UK

[52]BHF Centre for Cardiovascular Science, The Queen's Medical Research Institute, University of Edinburgh, UK

[53]Lancashire Teaching Hospital, UK

[54]School of Medicine, University of Nottingham, UK

[55]Leeds Teaching Hospitals NHS Trust, UK

[56]University of Birmingham, UK

[57]Liverpool University Hospitals Foundation NHS Trust, UK

[58]Salford Royal NHS Foundation Trust, UK

[59]Department of Clinical Genetics, Guy's and St Thomas' NHS Foundation Trust, UK

[60]Centre for Health and Related Research, School of Population Health, University of Sheffield, UK

[61]University College London Department of Renal Medicine, Royal Free Hospital, UK

[62]SW Thames Renal Unit, Epsom and St Helier University Hospitals NHS Trust, UK

[63]Alport UK, UK

[64]Queen Elizabeth University Hospital, Glasgow, UK

[65]College of Medicine and Health, University of Exeter, UK

[66]Divison of Population Health, University of Sheffield, UK

[67]University of Wolverhampton, UK

[68]Patient Representative, UK

[69]Institute of Liver Studies, King's College London, UK

[70]Sheffield Kidney Institute, Sheffield Teaching Hospitals NHS Foundation Trust, UK

[71]Royal Free Hospital, UK

[72]Manchester Institute of Nephrology and Transplantation, Manchester Royal Infirmary, UK

[73]PKD Charity, UK

[74]University Hospital Southampton NHS Foundation Trust, UK

[75]Children's Kidney centre, University Hospital of Wales, UK

[76]Travere Therapeutics, UK

[77]University Hospitals Coventry and Warwickshire NHS Trust, UK

[78]County Durham & Darlington NHS Foundation Trust, UK

[79]University of Leicester, UK

[80]Wirral University Teaching Hospital NHS Foundation Trust, UK

[81]University Hospitals Birmingham NHS Foundation Trust, UK

[82]Department of Medicine, University of Cambridge, UK

[83]North Bristol NHS Trust, UK

[84]Alder Hey Childrens NHS Foundation Trust, UK

[85]York & Scarborough Teaching Hospitals NHS Foundation Trust, UK

[86]Norfolk and Norwich University Hospitals NHS Trust, UK

[87]Royal Manchester Children's Hospital, Manchester, UK

[88]PTEN UK and Ireland Patient Group

[89]HNF1b Support Group, UK

[90]School of Medicine, Keele University, UK

[91]Wellcome Centre for Cell-Matrix Research, University of Manchester, UK

[92]Research Department of Pathology, University College London, UK

[93]HLRCC Foundation, UK

[94]Cambridge Institute of Therapeutic Immunology and Infectious Disease, Cambridge, UK

[95]Newcastle University, UK

[96]United Lincolnshire Hospitals NHS Trust, UK

[97]Department of Medical Genetics, University of Cambridge, UK

[98]University Hospitals of Derby and Burton NHS Foundation Trust, UK

[99]Retroperitoneal Fibrosis (RF) Group, UK

[100]National Institute of Health and Care Research Leeds Biomedical Research Centre, Leeds Teaching Hospitals NHS Trust, UK

[101]School of Medicine, University of Leeds, UK

[102]University of Liverpool, UK

[103]Newcastle Upon Tyne Hospitals NHS Foundation Trust, UK

[104]Research Department of Structural and Molecular Biology, University College London, UK

[105]Division of Cardiology, Cliniques Universitaires Saint-Luc, Belgium

[106]Pole of Cardiovascular Research, Institut de Recherche Expérimentale et Clinique, Université Catholique de Louvain, Brussels, Belgium

[107]Cambridge University Hospitals NHS Foundation Trust, UK

[108]East and North Hertfordshire NHS Trust, UK

[109]Department of Immunology and Inflammation, Faculty of Medicine, Imperial College London, UK

[110]Shrewsbury and Telford Hospital NHS Trust, UK

[111]National Institute of Health and Care Research Great Ormond Street Hospital Biomedical Research Centre, UK

[112]UCL Great Ormond Street Institute of Child Health, UK

[113]Northern Care Alliance NHS Foundation Trust, UK

[114]South Tyneside and Sunderland NHS Foundation Trust, UK

[115]Doncaster and Bassetlaw Teaching Hospitals, UK

[116]New Cross Hospital, Wolverhampton, UK

[117]Wellcome Centre for Mitochondrial Research, Translational & Clinical Research Institute, Faculty of Medical Sciences, Newcastle University, UK

[118]Great North Children's Hospital, Newcastle Upon Tyne, UK

[119]University of Edinburgh, UK

[120]Calderdale & Huddersfield Foundation Trust, UK

[121]Royal Surrey County Hospital, Guildford, UK

[122]South Tees Hospitals NHS Foundation Trust, UK

[123]University College Cork, Ireland

[124]National Amyloidosis Centre, University College London, UK

[125]Great Western Hospital, Swindon, UK

[126]North Tees and Hartlepool NHS Foundation Trust, UK

[127]University College London, UK

[128]aHUS Alliance, UK

[129]School of Biological Sciences, University of Manchester, UK
