## Supplementary appendix 2 Inclusion-Exclusion criteria for "The National Registry of Rare Kidney Diseases (RaDaR): Description, recruitment, and cross-sectional analyses of 25,880 adults and children with rare kidney diseases in the UK"

### RaDaR Inclusion and Exclusion Criteria

| Diagnosis | Cohort | Inclusion Criteria | Exclusion Criteria | Date of Diagnosis |
|---|---|---|---|---|
| **Adenine Phosphoribosyltransferase Deficiency (APRT-D)** | APRT Deficiency | APRT Deficiency confirmed<br><br>Abolished APRT enzyme activity or confirmed disease-causing mutation | None, if APRT Deficiency not confirmed | Date that clinical diagnosis was first made |
| **Alport Syndrome and Type IV collagenopathies** | Alport | Alport Syndrome definite or probable<br><br>Alport carrier definite or probable<br><br>Female heterozygote for X-linked Alport Syndrome (COL4A5)<br><br>Heterozygote for autosomal Alport Syndrome (COL4A3, COL4A4)<br><br>Thin basement membrane nephropathy | None stated | Date that clinical diagnosis was first made |
| **APOL1 disease, suspected or confirmed** | CKD-Africa Genes | People of African or Afro-Caribbean ancestry with CKD (KDIGO definition), >18 years<br><br>including: Focal segmental glomerulosclerosis (primary or secondary) on renal biopsy; Non-diabetic and non-immunological kidney disease with no other confirmed cause | None stated | Date that clinical diagnosis was first made |
| **Autoimmune distal renal tubular acidosis** | Tubulopathy | Autoimmune distal renal tubular acidosis | None stated | Date that clinical diagnosis was first made |
| **Autosomal dominant distal renal tubular acidosis** | Tubulopathy | Autosomal dominant distal renal tubular acidosis<br><br>Genetically confirmed heterozygous pathogenic variant in SLC4A1 | None stated | Date that clinical diagnosis was first made |
| **Autosomal recessive distal renal tubular acidosis** | Tubulopathy | Autosomal recessive distal renal tubular acidosis<br><br>Genetically confirmed homozygous pathogenic variant in ATP6V0A4, ATP6V1B1 or FOXI1 | None stated | Date that clinical diagnosis was first made |

| Diagnosis | Cohort | Inclusion Criteria | Exclusion Criteria | Date of Diagnosis |
|---|---|---|---|---|
| **Autosomal recessive proximal renal tubular acidosis** | Tubulopathy | Autosomal recessive proximal renal tubular acidosis with ocular abnormalities and intellectual disability<br><br>Genetically confirmed homozygous pathogenic variant in SLC4A4 | None stated | Date that clinical diagnosis was first made |

#### RaDaR Inclusion and Exclusion Criteria

| Diagnosis | Cohort | Inclusion Criteria | Exclusion Criteria | Date of Diagnosis |
|---|---|---|---|---|
| **Bartter Syndrome types 1 and 2** | Tubulopathy | Bartter Syndrome, infantile onset<br><br>Hypokalaemic alkalosis, infantile onset without hypertension<br><br>Hypokalaemic alkalosis, infantile onset with raised renin | Acidosis<br><br>Persistent Hyperkalaemia | Date that clinical diagnosis was first made |
| **Bartter Syndrome type 3 Gitelman Syndrome** | Tubulopathy | Bartter Syndrome type 3<br><br>Gitelman Syndrome<br><br>Hypokalaemic alkalosis with hypomagnesaemia<br><br>Hypokalaemic alkalosis with raised renin<br><br>Hypokalaemic alkalosis without hypertension | Acidosis<br><br>Hyperkalaemia | Date that clinical diagnosis was first made |
| **Bartter Syndrome Type 4** | Tubulopathy | Bartter Syndrome, infantile onset with deafness<br><br>Hypokalaemic alkalosis, infantile onset without hypertension with deafness<br><br>Hypokalaemic alkalosis, infantile onset with raised renin, with deafness | Acidosis<br><br>Persistent Hyperkalaemia | Date that clinical diagnosis was first made |
| **BK Nephropathy** | BK Nephropathy | Significant BK viraemia, with polymerase chain reaction (PCR) greater than or equal to 10 log 4 copies per ml.<br><br>A confirmatory biopsy is **not** required. | None stated | Date that PCR first equalled or exceeded 10 log 4 |

#### RaDaR Inclusion and Exclusion Criteria

| Diagnosis | Cohort | Inclusion Criteria | Exclusion Criteria | Date of Diagnosis |
|---|---|---|---|---|
| **Calciphylaxis** | Calciphylaxis | Any patient with a diagnosis of clinical diagnosis of Calciphylaxis; tissue diagnosis not required | None stated | Date that the diagnosis was made by a nephrologist or dermatologist |
| **Cystinosis (Nephropathic Cystinosis)** | Cystinosis | Cystinosis | None stated | Date that biochemical testing first showed an elevated level of white blood cell cysteine |
| **Cystinuria** | Cystinuria | Biochemically proven cystine kidney stone<br><br>Urinary cystine level > 3X reference range of the laboratory it was taken in<br><br>Cystine crystals in the urine (biochemically proven) | Another cause of proximal tubular dysfunction accounting for the raised cystine level e.g. Fanconi's syndrome | Date that any of the inclusion criteria first occurred |
| **Dent Disease** | Dent & Lowe | Dent Disease | None stated | Date that the clinical label of Dent Disease was first applied |
| **Dominant hypophosphatemia with nephrolithiasis or osteoporosis** | Tubulopathy | Dominant hypophosphatemia with nephrolithiasis or osteoporosis<br><br>Genetically confirmed heterozygous pathogenic variant in SLC34A1, SLC9A3R1, SLC34A3 | None stated | Date that clinical diagnosis was first made |
| **Drug induced Fanconi syndrome** | Tubulopathy | Drug induced Fanconi syndrome | None stated | Date that clinical diagnosis was first made |
| **Drug induced hypomagnesemia** | Tubulopathy | Drug induced hypomagnesemia | None stated | Date that clinical diagnosis was first made |

#### RaDaR Inclusion and Exclusion Criteria

| Diagnosis | Cohort | Inclusion Criteria | Exclusion Criteria | Date of Diagnosis |
|---|---|---|---|---|
| **Drug induced Nephrogenic Diabetes Insipidus** | Tubulopathy | Drug induced Nephrogenic Diabetes Insipidus | None stated | Date that clinical diagnosis was first made |
| **EAST syndrome (Epilepsy, Ataxia, Sensorineural deafness, Tubulopathy)** | Tubulopathy | Gitelman/Bartter-type syndrome in childhood with epilepsy /ataxia | Normal CNS examination | Date that clinical diagnosis was first made |
| **End stage kidney disease of unknown cause** | CKD-Africa Genes | People of African or Afro-Caribbean ancestry with CKD (KDIGO definition), >18 years | Known cause of kidney disease identified (unless Sickle cell Nephropathy or APOL1 disease) | Date that clinical diagnosis was first made |
| **Fabry Disease** | Fabry | Confirmed diagnosis of Fabry Disease | None stated | Date that genetic diagnosis was made and/or, for males, the date that low alpha gal levels were first recorded |
| **Familial Hypomagnesaemia with hypercalciuria and nephrocalcinosis CLDN16/19** | Tubulopathy | Familial Hypomagnesaemia with Hypercalciuria and Nephrocalcinosis<br><br>Genetically confirmed homozygous pathogenic variant in CLDN 16/19 | None stated | Date that clinical diagnosis was first made |
| **Familial primary hypomagnesemia with hypocalcuria FXYD2** | Tubulopathy | Familial primary hypomagnesemia with hypocalciuria<br>Genetically confirmed homozygous pathogenic variant in FXYD2 | None stated | Date that clinical diagnosis was first made |
| **Familial primary hypomagnesemia with normocalcuria EGF** | Tubulopathy | Familial primary hypomagnesemia with normocalcuria<br><br>Genetically confirmed homozygous pathogenic variant in EGF | None stated | Date that clinical diagnosis was first made |
| **Familial renal glucosuria** | Tubulopathy | Familial renal glucosuria | None stated | Date that clinical diagnosis |

### RaDaR Inclusion and Exclusion Criteria

| Diagnosis | Cohort | Inclusion Criteria | Exclusion Criteria | Date of Diagnosis |
|---|---|---|---|---|
| **SLC5A2** | | Genetically confirmed homozygous pathogenic variant in SLC5A2 | | was first made |

#### RaDaR Inclusion and Exclusion Criteria

| Diagnosis | Cohort | Inclusion Criteria | Exclusion Criteria | Date of Diagnosis |
|---|---|---|---|---|
| **Fanconi Renotubular syndrome 1 (FRTS1)** | Tubulopathy | Fanconi Renotubular syndrome 1 | None stated | Date that clinical diagnosis was first made |
| **Fanconi Renotubular syndrome 2 (FRTS2)** | Tubulopathy | Fanconi Renotubular syndrome 2<br><br>Genetically confirmed homozygous pathogenic variant in SLC34A1 | None stated | Date that clinical diagnosis was first made |
| **Fanconi Renotubular syndrome 3 (FRTS3)** | Tubulopathy | Fanconi Renotubular syndrome 3<br><br>Genetically confirmed homozygous pathogenic variant in EHHADH | None stated | Date that clinical diagnosis was first made |
| **Fibromuscular Dysplasia** | Fibromuscular Dysplasia | Diagnosis of FMD established on radiological or histological grounds<br><br>FMD of any arterial bed | None stated | Date that FMD was diagnosed by radiological (or histological) methods |
| **Generalized pseudohypoaldosteronism type 1** | Tubulopathy | Generalized pseudohypoaldosteronism type 1<br><br>Genetically confirmed homozygous pathogenic variant in SCNN1A/ SCNN1B/SCNN1G | None stated | Date that clinical diagnosis was first made |

#### RaDaR Inclusion and Exclusion Criteria

| Diagnosis | Cohort | Inclusion Criteria | Exclusion Criteria | Date of Diagnosis |
|---|---|---|---|---|
| **Haemolytic Uraemic Syndrome - Atypical** | aHUS | Diarrhoea-negative HUS, includes congenital and familial HUS<br><br>Renal biopsy showing a TMA and/or the triad of microangiopathic haemolytic anaemia, thrombocytopenia, renal failure. | Shiga toxin associated HUS<br><br>Secondary causes:<br>• Drugs<br>• Infection (HIV, pneumonia, streptococcus)<br>• Transplantation (bone marrow, liver, lung, cardiac but not de-novo renal)<br>• Cobalamin deficiency<br>• SLE<br>• APL Ab syndrome<br>• Scleroderma<br>• ADAMTS13 antibodies or deficiency | Date of first presentation |

#### RaDaR Inclusion and Exclusion Criteria

| Diagnosis | Cohort | Inclusion Criteria | Exclusion Criteria | Date of Diagnosis |
|---|---|---|---|---|
| **Haemolytic Uraemic Syndrome-Shiga toxin (Verocytotoxin)-associated** | STEC-HUS | Acute kidney injury (AKI) with elevated creatinine for age and/or oligoanuria (urine output <0.5ml/kg/hr over 24hr period) with either:<br>• Microangiopathic haemolytic anaemia (MAHA) - defined as Hgb < 10mg/dl with fragmented RBCs<br>or<br>• Thrombocytopaenia - defined as platelet count less than 130, 000 x 10 9/l<br>and<br>• Occurring with Shiga-toxin producing E Coli (STEC) infection defined as:<br>• Positive STEC culture<br>• Positive PCR for Stx gene directly from a faecal specimen<br>• Positive antibodies to the lipopolysaccharide<br>• antigen of E. coli serogroups O157, O26, O103, O111 and O145 | Septicaemia<br>Malignant hypertension<br>Primary vascular disease<br>Familial HUS not being part of the same | Date on which the STEC-HUS was suspected. |
| **Heavy metal induced Fanconi syndrome** | Tubulopathy | Heavy metal induced Fanconi syndrome | None stated | Date that clinical diagnosis was first made |

### RaDaR Inclusion and Exclusion Criteria

| Diagnosis | Cohort | Inclusion Criteria | Exclusion Criteria | Date of Diagnosis |
|---|---|---|---|---|
| **Hepatocyte Nuclear Factor-1B mutation** | HNF1b | Hepatocyte nuclear factor-1B mutation<br><br>Renal cysts and diabetes (RCAD)<br><br>Inherited genetic diabetes type 2 (MODY 5). | None stated | Date of genetic diagnosis |
| **Hereditary renal hypouricemia** | Tubulopathy | Hereditary renal hypouricemia<br><br>Genetically confirmed homozygous pathogenic variant in SLC22A12, SLC2A9 | None stated | Date of genetic diagnosis |
| **Hereditary hypophosphatemic rickets with hypercalciuria** | Tubulopathy | Hereditary hypophosphatemic rickets with hypercalciuria<br><br>Genetically confirmed homozygous pathogenic variant in SLC34A3 | None stated | Date of genetic diagnosis |
| **Hyperoxaluria (Primary hyperoxaluria, Oxalosis)** | Hyperoxaluria | Primary Hyperoxaluria Type1<br><br>Primary Hyperoxaluria Type 2<br><br>Primary Hyperoxaluria Type 3<br><br>Primary Hyperoxaluria awaiting genetic confirmation (Urine oxalate excretion ≥ 0.8 mmol/1.73 $m^2$/24 hrs)<br><br>Primary Hyperoxaluria Unclassified<br><br>Primary Hyperoxaluria Unclassified but with systemic oxalate deposition | Secondary hyperoxaluria associated with gastrointestinal disease<br><br>Renal failure without systemic oxalate deposits | Date that definitive diagnosis by genetic confirmation with gene mutation was first made.<br><br>If in doubt use the earliest date that PH was suspected or the date when treatment was first introduced |

#### RaDaR Inclusion and Exclusion Criteria

| Diagnosis | Cohort | Inclusion Criteria | Exclusion Criteria | Date of Diagnosis |
|---|---|---|---|---|
| **Hypertensive kidney disease** | CKD-Africa Genes | People of African or Afro-Caribbean ancestry with CKD (KDIGO definition), >18 years | Known cause of Kidney disease | Date that clinical diagnosis was first made |
| **Hyperuricaemic Nephropathy (Primary/Familial Hyperuricaemic nephropathy)**<br><br>**Medullary cystic kidney disease** | ADTKD | Autosomal Dominant Tubulointerstitial Kidney Disease (ADTKD; previously known as FUAN)<br><br>Familial juvenile hyperuricaemic nephropathy<br><br>Familial gouty nephropathy<br><br>Familial urate nephropathy<br><br>Familial interstitial nephropathy<br><br>Uromodulin-associated nephropathy<br><br>Medullary cystic kidney disease (type I or II) | None stated | Date that genetic confirmation was received |
| **IgA Nephropathy** | IgA Nephropathy | Biopsy proven IgA Nephropathy plus proteinuria >0.5g/ day or eGFR<60ml/min | All forms of secondary IgA nephropathy, including Henoch Schonlein purpura | Date of renal biopsy |
| **Isolated autosomal dominant hypomagnesemia, Glaudemans type** | Tubulopathy | Isolated autosomal dominant hypomagnesemia<br><br>Genetically confirmed homozygous pathogenic variant in KCNA1 | None stated | Date that clinical diagnosis was first made |

#### RaDaR Inclusion and Exclusion Criteria

| Diagnosis | Cohort | Inclusion Criteria | Exclusion Criteria | Date of Diagnosis |
|---|---|---|---|---|
| **Liddle syndrome** | Tubulopathy | Liddle syndrome<br><br>Hypertension with hypokalaemia, suppressed aldosterone<br><br>Hypertension with suppressed aldosterone<br><br>Autosomal dominant hypertension, suppressed aldosterone | Hyperaldosteronism | Date that clinical diagnosis was first made |
| **Lowe Syndrome** | Dent & Lowe | Lowe Syndrome | None Stated | Date that the clinical label of Lowe Syndrome was first applied |

#### RaDaR Inclusion and Exclusion Criteria

| Diagnosis | Cohort | Inclusion Criteria | Exclusion Criteria | Date of Diagnosis |
|---|---|---|---|---|
| **Membranoproliferative glomerulonephritis**<br><br>**Mesangiocapillary glomerulonephritis**<br><br>**Dense Deposit Disease**<br><br>**C3 Glomerulonephritis**<br><br>**C3 Glomerulopathy** | MPGN | Child or adult with histological finding of:<br><br>MPGN Type I<br><br>Dense Deposit Disease (morphological pattern may or may not be MPGN)<br><br>Other pattern of MPGN<br><br>C3 Glomerulonephritis (Characterised by C3 deposits in the absence of immunoglobulin with electron dense deposits (morphological pattern may or may not be MPGN)<br><br>Unclassified GN with capillary wall immune deposits | MPGN known to be secondary to:<br><br>Chronic bacterial infection<br><br>Hepatitis B or C infection<br><br>Malignancy<br><br>Systemic lupus erythematosus (by ACR criteria) | Date of biopsy |
| **Membranous Nephropathy** | Membranous Nephropathy | Membranous nephropathy confirmed by kidney histology | Lupus nephritis | Date of biopsy |
| **Mitochondrial Renal Disease** | Mitochondrial | Mitochondrial Disease **or** Mitochondrial Cytopathy | None Stated | Date that clinical diagnosis was first made |

### RaDaR Inclusion and Exclusion Criteria

| Diagnosis | Cohort | Inclusion Criteria | Exclusion Criteria | Date of Diagnosis |
|---|---|---|---|---|
| **Monoclonal Gammopathy of Renal Significance** | MGRS | Renal biopsy proven confirmation of:<br><br>• AH amyloidosis*<br>• AHL amyloidosis*<br>• AL amyloidosis*<br>• C3 glomerulonephritis with monoclonal gammopathy<br>• Crystalglobulinaemia<br>• Crystal-storing histiocytosis<br>• Fibrillary Glomerulonephritis<br>• Immunotactoid/Glomerulonephritis with Organised Microtubular Monoclonal Immunoglobulin Deposits (GOMMID)<br>• Intracapillary monoclonal IgM without cryoglobulin<br>• Intraglomerular/capillary lymphoma/leukaemia<br>• Light chain cast nephropathy<br>• Light chain proximal tubulopathy, crystalline<br>• Light chain proximal tubulopathy, non crystalline<br>• Monoclonal Immunoglobulin Deposition Disease (MIDD; includes Light Chain Deposition Disease - LCDD; Heavy Chair Deposition Disease - HCDD; and Light and Heavy Chain Deposition Disease - LHCDD)<br>• Proliferative glomerulonephritis with monoclonal immunoglobulin deposits – PGNMID<br>• Thrombotic Microangiopathy with monoclonal gammopathy<br>• Type 1 cryoglobulinaemic Glomerulonephritis<br>• Unclassified MGRS<br><br>*Patients with systemic amyloidosis may have a renal biopsy confirming AL amyloidosis or a biopsy of other tissue with confirmation of renal involvement by the UK National Amyloidosis Centre. | None Stated | Date of biopsy |

#### RaDaR Inclusion and Exclusion Criteria

| Diagnosis | Cohort | Inclusion Criteria | Exclusion Criteria | Date of Diagnosis |
|---|---|---|---|---|
| **Nephrogenic diabetes insipidus** | Tubulopathy | Nephrogenic diabetes insipidus<br><br>Genetically confirmed homozygous pathogenic variant in AVPR2, AQP2 | None stated | Date that clinical diagnosis was first made |
| **Nephrogenic syndrome of inappropriate antidiuresis** | Tubulopathy | Nephrogenic syndrome of inappropriate antidiuresis<br><br>Genetically confirmed homozygous pathogenic variant in AVPR2 | None stated | Date that clinical diagnosis was first made |
| **Nephronophthisis** | ARPKD/NPHP | Histological or radiological features of Nephronophthisis<br><br>Genetic diagnosis of Nephronophthisis or Nephronophthisis-related ciliopathy | None stated | Date that histological /radiological or genetic diagnosis was first made |

#### RaDaR Inclusion and Exclusion Criteria

| Diagnosis | Cohort | Inclusion Criteria | Exclusion Criteria | Date of Diagnosis |
|---|---|---|---|---|
| **Nephrotic Syndrome - Steroid Sensitive or Steroid Resistant**<br><br>**(Congenital nephrotic syndrome, nephrotic syndrome with focal segmental glomerulosclerosis)** | INS | Children and adults with idiopathic Nephrotic Syndrome (nephrotic range proteinuria and hypoalbuminaemia)<br><br>Congenital NS (presumed Steroid Resistance)<br><br>Childhood or adult onset with primary Steroid Resistance<br><br>Childhood or adult onset with late onset Steroid Resistance<br><br>Steroid Sensitive Nephrotic Syndrome (full or partial remission in response to steroids)<br><br>As part of a syndrome e.g. Nail Patella Syndrome and Denys-Drash Syndrome<br><br>Those with a biopsy diagnosis of FSGS or minimal change disease can be included if they fall in the above categories but biopsy is not a prerequisite for inclusion | Secondary causes of Nephrotic Syndrome<br><br>• Primary diagnosis of Glomerulonephritis (IgA Nephropathy, Membranoproliferative Glomerulonephritis, Membranous Nephropathy)<br>• Vasculitis<br>• Systemic Lupus Erythematosus<br>• Diabetes<br>• Obesity<br>• Hypertension | Date of presentation to secondary or tertiary centre |
| **Oncogenic osteomalacia** | Tubulopathy | Oncogenic osteomalacia | None stated | Date that clinical diagnosis was first made |
| **Osteopetrosis with renal tubular acidosis** | Tubulopathy | Osteopetrosis with renal tubular acidosis<br><br>Genetically confirmed homozygous pathogenic variant in CA2 | None stated | Date that clinical diagnosis was first made |

#### RaDaR Inclusion and Exclusion Criteria

| Diagnosis | Cohort | Inclusion Criteria | Exclusion Criteria | Date of Diagnosis |
|---|---|---|---|---|
| **Polycystic Kidney Disease - Autosomal Dominant** | ADPKD | Clinical features of Autosomal Dominant Polycystic Kidney Disease meeting current image based diagnostic criteria<br><br>Clinical features compatible with ADPKD in the absence of a family history<br><br>Pathogenic or likely pathogenic PKD1 or PKD2 mutation with or without clinical features | Autosomal dominant polycystic liver disease with no evidence of renal cysts | Date that the clinical diagnosis was first made.<br><br>This may be reported by the clinician as the date of the diagnostic scan or by the patient if scans were performed at another centre |
| **Polycystic Kidney Disease - Autosomal Recessive** | ARPKD | Autosomal Recessive Polycystic Kidney Disease<br><br>Congenital Hepatic Fibrosis<br><br>Caroli Syndrome with kidney malformation or cyst | None stated | Date that clinical diagnosis was first made. |

#### RaDaR Inclusion and Exclusion Criteria

| Diagnosis | Cohort | Inclusion Criteria | Exclusion Criteria | Date of Diagnosis |
|---|---|---|---|---|
| **Pregnancy and Chronic Kidney Disease** | Pregnancy | Pregnancy in all women known to have CKD 1-5 prior to pregnancy or those with a serum creatinine >85umol/l on two occasions during pregnancy<br><br>Pregnancy in all women with renal transplants regardless of function<br><br>Pregnancy in all women with previous or current lupus nephritis regardless of function | None stated | Date of last menstrual period |
| **Primary hypomagnesemia with secondary hypocalcemia** | Tubulopathy | Primary hypomagnesemia with secondary hypocalcemia<br><br>Genetically confirmed homozygous pathogenic variant in TRPM6 | None stated | Date that clinical diagnosis was first made |
| **Pseudohypoaldosteronism type 2A** | Tubulopathy | Pseudohypoaldosteronism type 2A | None stated | Date that clinical diagnosis was first made |
| **Pseudohypoaldosteronism type 2B** | Tubulopathy | Pseudohypoaldosteronism type 2B<br><br>Genetically confirmed homozygous pathogenic variant in WNK1 | None stated | Date that clinical diagnosis was first made |
| **Pseudohypoaldosteronism type 2C** | Tubulopathy | Pseudohypoaldosteronism type 2C<br><br>Genetically confirmed homozygous pathogenic variant in WNK4 | None stated | Date that clinical diagnosis was first made |
| **Pseudohypoaldosteronism type 2D** | Tubulopathy | Pseudohypoaldosteronism type 2D<br><br>Genetically confirmed homozygous pathogenic variant in KLHL3 | None stated | Date that clinical diagnosis was first made |

#### RaDaR Inclusion and Exclusion Criteria

| Diagnosis | Cohort | Inclusion Criteria | Exclusion Criteria | Date of Diagnosis |
|---|---|---|---|---|
| **Pseudohypoaldosteronism type 2E** | Tubulopathy | Pseudohypoaldosteronism type 2E<br><br>Genetically confirmed homozygous pathogenic variant in CUL3 | None stated | Date that clinical diagnosis was first made |
| **Pure Red Cell Aplasia** | PRCA | Treatment with any injectable form of erythropoiesis stimulating agent for at least four weeks.<br><br>Haemoglobin <70 g/l without transfusion or transfusion dependence.<br><br>Normal leucocyte and platelet count<br><br>Reticulocyte count < 20.000 / mm3<br><br>Bone marrow aspirate showing well preserved myeloid and megakaryocyte development, and <5% erythroblasts.<br><br>Presence of anti-erythropoietin antibodies. | Pre-established PRCA due to myeloproliferative disorder | Date of positive antibody test |
| **Renal pseudohypoaldosteronism type 1** | Tubulopathy | Renal pseudohypoaldosteronism type 1<br><br>Genetically confirmed homozygous pathogenic variant in NR3C2 | | Date that clinical diagnosis was first made |

#### RaDaR Inclusion and Exclusion Criteria

| Diagnosis | Cohort | Inclusion Criteria | Exclusion Criteria | Date of Diagnosis |
|---|---|---|---|---|
| **Retroperitoneal Fibrosis** | Retroperitoneal Fibrosis | Any radiologically confirmed retroperitoneal fibrosis (RPF), presumed to be 'idiopathic' or associated with primary conditions including (but not exclusively):<br>• Aortitis<br>• Periaortitis<br>• IgG4-related Vasculitis<br>• Perivascular fibrosis<br>• Atherosclerotic or aneurysmal disease<br><br>**Note:** There is no specific ICD code for retroperitoneal fibrosis although the diagnosis term links to two ICD codes:<br>• ICD10:N13.5 - Crossing vessel and stricture of ureter without hydronephrosis<br>• ICD-9-CM 593.4 - Other ureteric obstruction | Neoplastic disease within retroperitoneal fibrosis mass defined histologically | Date of diagnostic imaging study report |
| **Sickle Cell Nephropathy** | **CKD Africa Genes** | People of African or Afro-Caribbean ancestry with CKD (KDIGO definition), >18 years<br><br>Known Sickle Cell disease with reduced kidney function, and/or blood or protein in urine with no other cause for kidney disease identified | None stated | Date that clinical diagnosis was first made |

| | | | | |
|---|---|---|---|---|
| **Tuberous Sclerosis** | **Tuberous Sclerosis** | Clinical or molecular diagnosis of Tuberous Sclerosis Complex (TSC)<br><br>Multiple renal angiomyolipomas<br><br>Multiple renal angiomyolipomas (> 3) +/- pulmonary lymphangioleiomyomatosis (LAM) without other signs of TSC | None stated | Date that clinical diagnosis was first made |

| | | | | |
|---|---|---|---|---|
| **Vasculitis (Primary systemic Vasculitis)** | **Vasculitis** | **Small vessel Vasculitis (ANCA associated)**<br><br>Microscopic polyangiitis (including renal limited Vasculitis)<br><br>Granulomatosis with polyangiitis (Wegener)<br><br>Eosinophilic granulomatosis with polyangiitis (Churg Strauss)<br><br>ANCA Vasculitis unclassified<br><br>**Small vessel Vasculitis (Immune complex)**<br><br>anti-GBM disease<br><br>Cryoglobulinemic Vasculitis<br><br>IgA Vasculitis (Henoch-Schönlein)<br><br>**Medium vessel Vasculitis**<br><br>Classical PAN<br><br>Kawasaki disease<br><br>**Large vessel Vasculitis**<br><br>Giant cell arteritis<br><br>Takayasu's arteritis | None stated | Date of biopsy.<br><br>In the absence of a biopsy, the date of a positive antibody test should be used |

| | | | | |
|---|---|---|---|---|
| **Vasculitis (Primary systemic Vasculitis)** | Vasculitis | **Variable vessel Vasculitis**<br><br>Behçet's disease<br><br>Cogan's syndrome<br><br>**Single organ Vasculitis**<br><br>Isolated aortitis<br><br>Primary cerebral angiitis | None stated | Date of biopsy.<br><br>In the absence of a biopsy, the date of a positive antibody test should be used |

| | | | | |
|---|---|---|---|---|
| **Inherited Renal Cancer Syndrome** | Renal Cancer Inherited | 1. A molecular or clinical diagnosis according to standard criteria of any of the following conditions:<br><br>Von Hippel Lindau disease (VHL) OMIM 193300<br><br>PTEN hamartoma tumour syndrome (Cowden syndrome) OMIM 158350<br><br>Birt Hogg Dube syndrome (BHD) OMIM 135150<br><br>Hereditary leiomyomatosis and renal cell cancer syndrome(HLRCC) OMIM 150800<br><br>Succinate dehydrogenase-related tumour predisposition syndrome<br><br>BAP1-related tumour predisposition syndrome OMIM 614327<br><br>Hereditary Type 1 papillary renal cell carcinoma syndrome (MET oncogene) OMIM 605074<br><br>2. Two or more cases in first degree relatives of any type of renal cancer without an established molecular or clinical diagnosis<br>**3.** Bilateral, multiple primary renal cancers of any histopathological type with or with a family history | None stated | Date of molecular or clinical diagnosis according to standard criteria |
